# Coupling machine learning and high throughput multiplex digital PCR enables accurate detection of carbapenem-resistant genes in clinical isolates

**DOI:** 10.1101/2021.04.16.21255464

**Authors:** Luca Miglietta, Ahmad Moniri, Ivana Pennisi, Kenny Malpartida-Cardenas, Hala Abbas, Kerri Hill-Cawthorne, Frances Bolt, Elita Jauneikaite, Frances Davies, Alison Holmes, Pantelis Georgiou, Jesus Rodriguez-Manzano

**Author notes:** **Correspondence to:** Jesus Rodriguez-Manzano, PhD, Department of Infectious Disease, Imperial College London, Room 8N9, Commonwealth Building, Hammersmith Hospital Campus, Du Cane Road, London, W12 0NN.

## Abstract

Rapid and accurate identification of patients colonised with carbapenemase-producing organisms (CPOs) is essential to adopt prompt prevention measures to reduce the risk of transmission. Recent studies have demonstrated the ability to combine machine learning (ML) algorithms with real-time digital PCR (dPCR) instruments to increase classification accuracy of multiplex PCR assays when using synthetic DNA templates. We sought to determine if this novel methodology could be applied to improve identification of the five major carbapenem-resistant genes in clinical CPO-isolates, which would represent a leap forward in the use of PCR-based data-driven diagnostics for clinical applications.

We collected 253 clinical isolates (including 221 CPO-positive samples) and developed a novel 5-plex PCR assay for detection of *bla*_IMP_, *bla*_KPC_, *bla*_NDM_, *bla*_OXA-48_ and *bla*_VIM_. Combining the recently reported ML method ‘Amplification and Melting Curve Analysis’ (AMCA) with the abovementioned multiplex assay, we assessed the performance of the AMCA methodology in detecting these genes. The improved classification accuracy of AMCA relies on the usage of real-time data from a single-fluorescent channel and benefits from the kinetic/thermodynamic information encoded in the thousands of amplification events produced by high throughput real-time dPCR.

The 5-plex showed a lower limit of detection of 10 DNA copies per reaction for each primer set and no cross-reactivity with other carbapenemase genes. The AMCA classifier demonstrated excellent predictive performance with 99.6% (CI 97.8-99.9%) accuracy (only one misclassified sample out of the 253, with a total of 160,041 positive amplification events), which represents a 7.9% increase (p-value < 0.05) compared to conventional melting curve analysis.

This work demonstrates the use of the AMCA method to increase the throughput and performance of state-of-the-art molecular diagnostic platforms, without hardware modifications and additional costs, thus potentially providing substantial clinical utility on screening patients for CPO carriage.

## 1 Introduction

This paper demonstrates that machine learning (ML) approaches coupled with high throughput real-time digital PCR (dPCR) can be used to increase detection accuracy of multiplex PCR assays when screening clinical isolates for the presence of carbapenemase-producing organisms (CPOs). We used a recently reported ML method called Amplification and Melting Curve Analysis (AMCA), which leverages the target-specific information encoded in each amplification event (via real-time data), to identify the nature of nucleic acid molecules (Moniri et al., 2020a). The AMCA approach is based on training supervised machine learning algorithms to extract kinetic and thermodynamic information from PCR amplification and melting curves to enhance the classification accuracy in multiplexing. Validation of this methodology using clinical isolates has never been reported before; therefore, this work represents a step forward towards the implementation of this method into clinical microbiology laboratories. Nucleic acid amplification tests (NAATs) that incorporate the AMCA classifier for multiple target detection will greatly improve their specificity, sensitivity and turn-around time to result, reducing overall resource consumptions and improving diagnostic performance.

Antimicrobial resistance (AMR) is a serious global threat and poses a challenge for modern medicine, compromising effective infectious disease management (Bush and Fisher, 2011; Tzouvelekis et al., 2012). One of the most concerning forms of AMR is the rapid spread of CPOs; bacteria producing enzymes that inactivate the potent antibiotics, carbapenems. Whilst overall UK incidence is low, there are centres nationally facing increasing rates and outbreaks, including Imperial College Healthcare NHS Trust (ICHNT), and it is endemic in many other regions worldwide (Otter et al., 2017b; Rodriguez-Manzano et al., 2020). CPO infections are associated with higher morbidity and mortality than susceptible strains, in part because their resistance can lead to ineffective empirical therapy and suboptimal treatment (Neuner et al., 2011; Eliopoulos et al., 2014). Therapeutic options are severely restricted, and in many cases clinical management relies on “last line” antibiotics that are less effective and have more side effects (Bleumin et al., 2012).

Patients infected with CPOs present significant challenges for diagnostics and infection control. There is an urgent need for accurate and timely diagnosis to improve patient outcomes and prevent the spread of AMR. Carbapenemase resistance genes are often co-localised on highly transmissible plasmids and are readily shared between bacterial species, providing the ideal conditions for multidrug resistant organisms (Johnning et al., 2018). Incorrect diagnosis delays appropriate intervention, increases financial burdens for the healthcare system, and complicates antimicrobial stewardship efforts (Charani et al., 2021). A local ICHNT economic analysis estimated the cost of a large hospital outbreak (∼100 infections) of carbapenemase producing *Klebsiella pneumoniae* to be £1M. Some of the increased expenditure was associated with increased screening, bed closures, medication and patient bed-days (Otter et al., 2017a); better diagnostics could reduce these costs.

Diagnosis of CPOs is often too complicated and time-consuming, as it is normally based upon multiple tests which employ a wide range of instruments and diagnostic tests. Phenotypic methods typically target carbapenemase production and provide no information on the underlying resistance mechanism (Codjoe and Donkor, 2017). These tests represent a low-cost (£2-15 per sample) and robust methodology; however, they rely on pure culture which increases turnaround times (12-24h) (Moloney et al., 2019). A variety of molecular methods, including amplification (PCR-based), microarray and sequencing assays have been developed and are frequently used in microbiology laboratories (Matsumura and Pitout, 2016; Reta et al., 2020). Microarray and sequencing are time consuming (>12-48h), expensive (>£50K platforms and >£80 per sample), and require bioinformatic expertise. Conversely, NAATs are commonly cheaper (£15-30 per sample) and faster (1-2h), whereas instrument price significantly ranges between tens to hundreds of thousands of pounds for conventional and digital PCR platforms, respectively (Huggett et al., 2015; Quan et al., 2018). Furthermore, the application of sophisticated data processing for its optimisation (as done with microarray and sequencing methods) has been largely unexplored (Collins and Moons, 2019; Beinhauerova et al., 2020). As a result of all aforementioned limitations, implementation of microarrays, sequencing and molecular methods for CPO diagnosis into routine practice is often limited.

Recently, our group has demonstrated that the large volume of data obtained from real-time digital PCR (dPCR) instruments can be exploited to perform data-driven multiplexing in a single fluorescent channel, reporting a 99.33 ± 0.13% classification accuracy when using synthetic DNA in a 9-plex format (Moniri et al., 2020a). This result represented an increase of 10% over using melting curve analysis, indicative of the potential benefits of this methodology for diagnostic and screening applications. The ML method used (AMCA) leverages kinetic and thermodynamic information encoded in the amplification and melting curves to perform target identification in multiplexed environments (Moniri et al., 2019, 2020b; Rodriguez-Manzano et al., 2019). Here we evaluate, for the first time, the analytical performance of AMCA method compared to Xpert Carba-R Cepheid and Resist-3 O.K.N assays when tested on clinical isolates for detection of the most common types of serine-beta-lactamases (*bla*_KPC_ and *bla*_OXA-48_) and metallo-beta-lactamases (*bla*_IMP_, *bla*_VIM_ and *bla*_NDM_) (Maurer et al., 2015; Lim et al., 2018). Results were compared against another ML based classifier ‘Melting Curve Analysis’ (MCA), which uses the thermodynamic information contained in PCR melting curves for identification of multiple targets in a single well reaction (Athamanolap et al., 2014; Moniri et al., 2020a). A 5-plex PCR assay was developed in-silico and validated with synthetic DNA templates. The performance of the AMCA method, using this 5-plex, was further assessed with 253 clinical isolates provided by the microbiology department at Charing Cross Hospital, ICHNT. All samples were analysed in real-time dPCR, using an intercalating dye (EvaGreen) in a single-fluorescent channel. This work demonstrates that the AMCA method can be integrated with conventional clinical diagnostic workflows in combination with real-time dPCR platforms, as it does not require any hardware modification. Increasing multiplexing capabilities enables improved workflow efficiency while reducing per sample cost, and it is beneficial to a number of application fields beyond clinical diagnostics, such as veterinary and environmental fields, where multiple targets need to be analysed simultaneously (e.g., SNP genotyping, forensic studies and gene deletion analysis). Figure 1 illustrates the concept of data-driven multiplexing, where tailored PCR-based amplification chemistries combined with advance data analytics can be seamlessly integrated into existing diagnostics pipelines which utilize real-time platforms.

**Figure 1.**
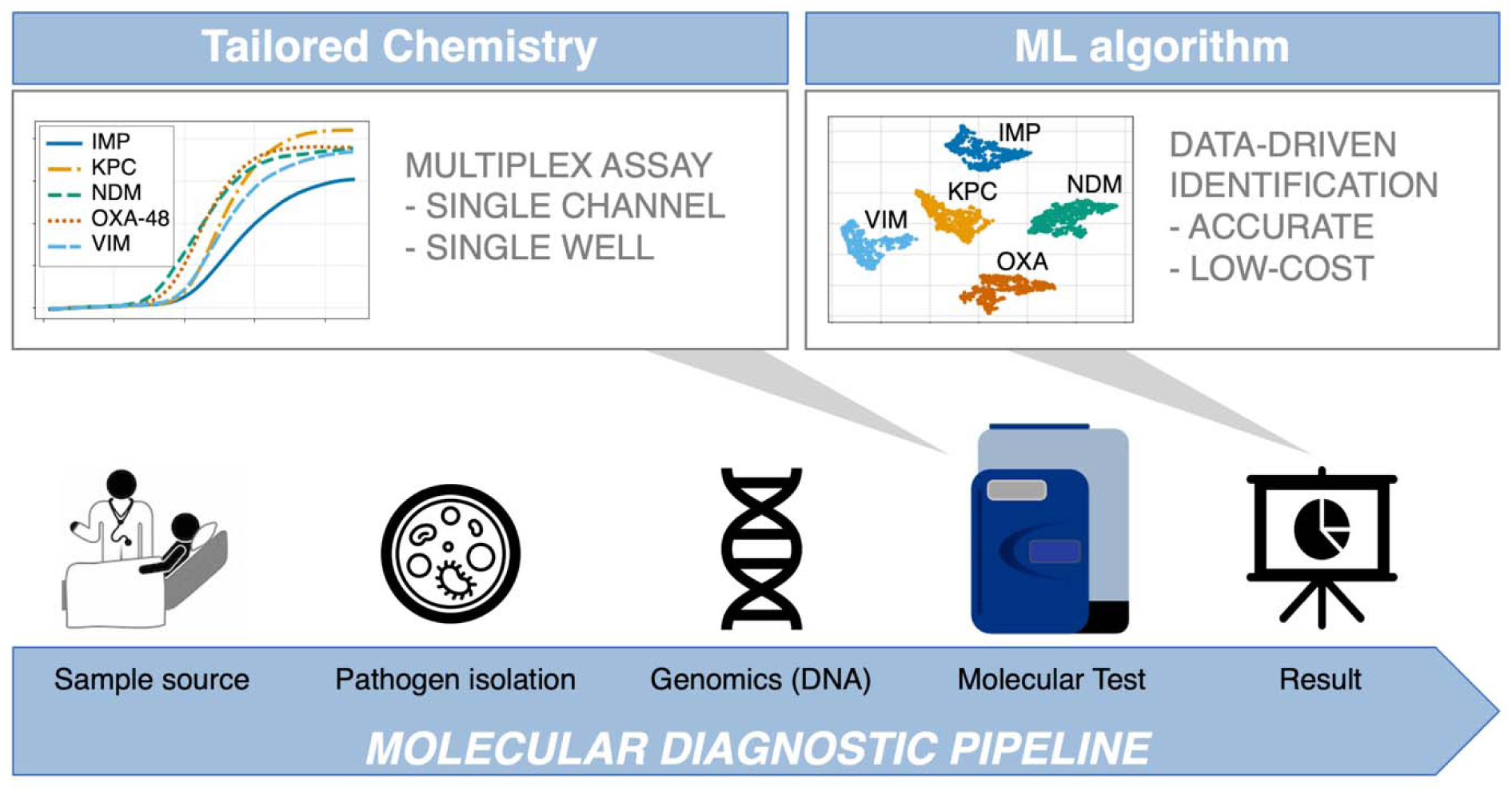
Integration of data-driven approaches to standard diagnostic workflows. The blue arrow indicates the conventional diagnosis pipeline from patient to result, where patient sample is collected from different sources (e.g., eye swab, nasopharyngeal swab, throat swab, urine, or rectal swab). Subsequently, samples are cultured, and nucleic acids are extracted in a microbiology lab. Following this, the most suitable genetic test is developed in-silico, comprising of specialised assays capable of multi target detection in a single reaction (first grey arrow). The test is performed in the dPCR instrument, outputting large amounts of data, which are analysed by a machine learning supported algorithm to ensure reliable and accurate results (second grey arrow). This is where the AMCA methodology is applied.

## 2 Experimental Section

### 2.1 Synthetic DNA

Double-stranded synthetic DNA (gBlocks^®^ Gene Fragments) containing the entire coding sequences of *bla*_IMP_, *bla*_KPC_, *bla*_NDM_, *bla*_OXA-48_ and *bla*_VIM_ genes was used for quantitative real-time PCR (qPCR) experiments when determining the limit-of-detection of the 5-plex PCR assay, and in dPCR experiments for generating the digital bulk standards and training the mathematical models. The gene fragments (ranging from 900 to 1000 bp) were purchased from Integrated DNA Technologies Ltd (IDT) and resuspended in Tris-EDTA buffer to 10 ng/μL stock solutions (stored at −80 °C until further use). The DNA stock concentration for all targets was estimated by dPCR using the Fluidigm’s Biomark HD system. The following NCBI accession numbers are used as reference for the gBlock synthesis: NG_049172 (*bla*_IMP_), NC_016846 (*bla*_KPC_), NC_023908 (*bla*_NDM_), NG_049762 (*bla*_OXA-48_) and NG_050336 (*bla*_VIM_).

### 2.2 Clinical isolates – bacterial strains and culture condition

A total of 253 non-duplicated *Enterobacteriaceae* isolates were collected between 2012-2020 from clinical or screening samples routinely processed by Microbiology Department at Charing Cross Hospital, ICHNT (Ethics protocol 06/Q0406/20). Species identification was performed using MALDI-TOF MS and carbapenemase mechanisms were determined using the Xpert Carba-R (Cepheid) or Resist-3 O.K.N assay (Corisbio). The isolates were subcultured on appropriate growth media and incubated at 37 °C overnight, and the genomic DNA was extracted using GenElute Bacterial Genomic DNA kit (Sigma-Aldrich) following the manufacturer’s instructions.

### 2.3 Primer Design

The genes used in this study belong to (i) class A carbapenemase encoding for *bla*_KPC_ type, (ii) class D oxacillinases encoding *bla*_OXA-48_ and (iii) class B metalloenzymes encoding *bla*_NDM_, *bla*_IMP_ and *bla*_VIM_. The sequences of these genes were downloaded from the GenBank website (http://www.ncbi.nlm.nih.gov/genbank/). Based on the comprehensive analyses and alignments of each carbapenemase type using the MUSCLE algorithm, primers were specifically designed to amplify all alleles of each carbapenemase gene family described above (Edgar, 2004). Design and in-silico analysis were conducted using GENEious Prime 2020.1.2 (https://www.geneious.com). Primer characteristics were analysed through IDT OligoAnalyzer software (https://eu.idtdna.com/pages/tools/oligoanalyzer) using the J. SantaLucia thermodynamic table for melting temperature (T_m_) evaluation, hairpin, self-dimer, and cross-primer formation (multiple-primer-analyzer @ www.thermofisher.com). The T_m_ of the amplification product of each gene was determined by Melting Curve Predictions Software (uMELT) package (https://dna-utah.org/umelt/umelt.html). To confirm the specificity of the real-time digital PCR assays, the primers were first evaluated in a singleplex PCR environment to ensure that they correctly amplified their respective loci and that the amplicons showed the predicted T_m_ and after that in multiplex format. All primers were synthesised by IDT (Coralville, IA, USA). Primer sequences and amplicon information are listed in Table 1.

**Table 1.**
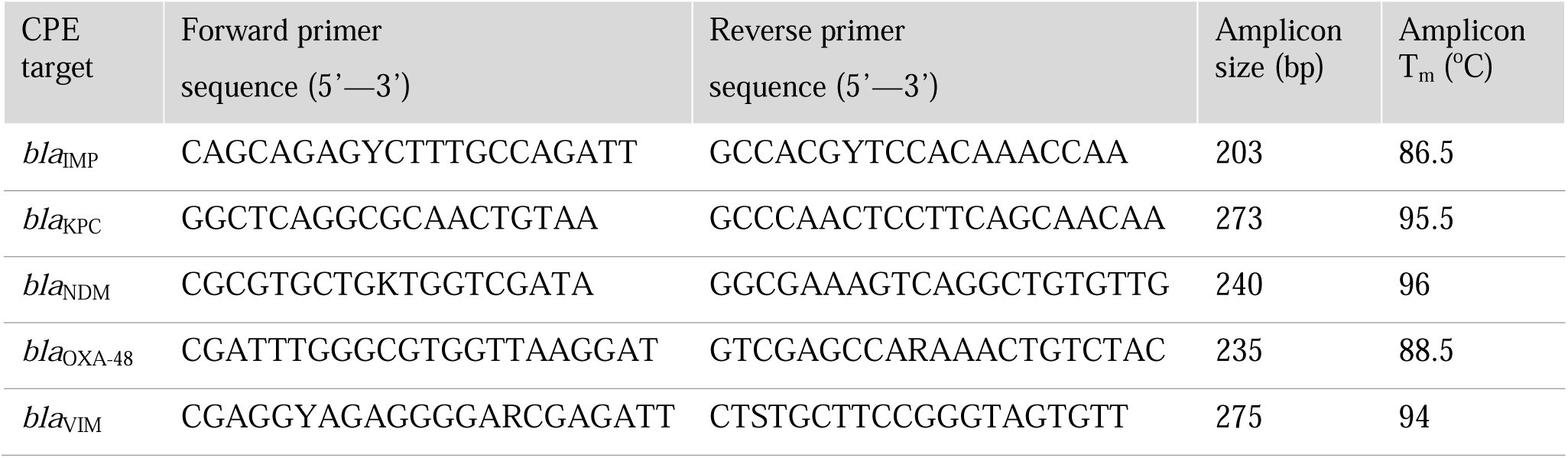
Primer sets developed in this study for the 5-plex PCR assay.

### 2.4 Multiplex real-time digital PCR

Each amplification mix for dPCR experiments contained the following: 2 μL of SsoFast EvaGreen Supermix with Low ROX (BioRad, UK), 0.4 μL of 20X GE Sample Loading Reagent (Fluidigm PN 85000746), 0.2 μL of PCR grade water, 0.2 μL of 20X multiplex PCR primer mixture containing the five primer sets (10 μM of each primer), and 1.2 μL of different concentrations of synthetic DNA, samples or controls to bring the final volume to 4 μL. PCR cycling condition consisted of a hot start step for 10 min at 95°C, followed by 45 cycles at 95°C for 20 s, 67°C for 45 s, and 72°C for 30 s. Melting curve analysis was performed with one cycle at 65°C for 3 s and reading from 65 to 97°C with an increment of 0.5°C. We used the integrated fluidic circuit controller to prime and load qdPCR 37K digital chips and Fluidigm’s Biomark HD system to perform the dPCR experiments, following manufacturer’s instructions. Each digital chip contains 48 inlets, where each inlet is connected to a microfluidic panel consisting of 770 partitions or wells (0.85 nL well volume). In this study, we used a total of 7 qdPCR 37K digital chips, totalling 336 panels and 189,206 positive amplification reactions (29,165 from training and 160,041 from testing experiments).

### 2.5 Limit of detection for the 5-plex PCR assay

Analytical sensitivity was evaluated with 10-fold dilutions of gBlocks^®^ Gene Fragments containing the sequence for the 5 carbapenemase genes, ranging from 10^1^ to 10^6^ DNA copies per reaction. Each experimental condition was run in triplicate. The qPCR assays were performed in a LightCycler 96 and the data was analysed using LC96 System software version SW1.1. Further details in the experimental conditions used for qPCR are provided in Supplementary Data S1.

### 2.6 Quantification of clinical isolates

Clinical isolates were quantified by real-time dPCR following the methodology proposed by Moniri et al 2020b. Thus, using Poisson statistics when the microfluidic panel occupancy was ≤ 85% (a maximum of 665 positive amplification events for a given panel) and quantification cycle (C_q_) interpolation from digital bulk standards when panel occupancy was >85%. Digital bulk standards were generated by serial dilutions of the gBlocks^®^ Gene Fragments containing the sequence for the 5 carbapenemase genes ranging from reaction 10^1^ to 10^5^ DNA copies per panel. The C_q_ values are calculated by the Fluidigm Digital PCR Analysis software 2.1.1.

### 2.7 Machine learning-based methods

The proposed method, AMCA, trains a supervised machine learning model in which the best fit linear line and the optimal value of intercept and coefficient are calculated to minimize error when combining the predictions of amplification curve analysis (ACA) and MCA, as previously reported in Moniri et al (Moniri et al., 2020a, 2020b). In this study, the ACA consists of applying a k-nearest neighbors (KNN) model (with parameter k=10) to the entire real-time curve from each amplification event, whereas the MCA method consists of applying a logistic regression model to T_m_ values extracted from each melting curve (Cunningham and Delany, 2020). Both ACA and MCA output 5 probabilities associated with each target in the 5-plex. Therefore, as showed in the flowchart in the Supplementary Figure S1, these probabilities are concatenated into 10 values which are the input to the AMCA method. It is important to note that this classifier is tuned with its own cross-validation step to avoid overfitting. The classifier threshold for positive samples has been set at 5% of panel occupancy, Further details of the AMCA linear regression model are described in Supplementary Data S2.

### 2.8 Statistical Analysis

(i) Sample size: A sufficient number of samples was determined to provide statistically significant results via the binomial proportion confidence interval method (Mercaldo ND, Lau KF, 2007). Under the assumption that the test has a sensitivity and specificity of 95% with a 5% margin of error, the number of samples were determined as 72 (which is significantly smaller than 221 used in this study). (ii) AMCA cross-validation performance: Prior to evaluating the in-sample performance of the model, by using the 221 clinical isolates, the out-of-sample classification accuracy was estimated by 10-fold cross-validation on the training data (using stratified splits). (iii) AMCA accuracy: The two-sided t-test with unknown variances was used to determine statistical significance for comparing the classification accuracy of AMCA against MCA. Prior to this test, a Lilliefors test was used to determine normality of the distributions and the Bartlett test for equal/unequal variances. A *p*-value of 0.05 was used as a threshold for statistical significance for all tests.

## 3 Results

### 3.1 Primer characterisation for optimal multiplex PCR assay performance

#### 3.1.1 in-silico analysis

To test the inclusivity and exclusivity of the 5-plex PCR assay, primers were subjected to a general NCBI BlastN search against more than 500 sequences per target. Inclusivity results showed over 99% identity coverage for each target (inclusivity alignments are provided in Supplementary Figure S2-S6). For exclusivity analysis, BlastN hits with an identity score lower than 80% were regarded as negative. No cross-reactivity was observed with other sequences deposited in the database.

#### 3.1.2 Experimental results in qPCR

The 5-plex PCR assay has been validated using a conventional qPCR platform with synthetic DNA templates at concentrations ranging from 10^1^ to 10^6^ DNA copies/reaction. Supplementary Figure S7 shows the real-time amplification, melting and standard curves obtained from analytical sensitivity experiments. The amplification and melting curves have distinct shape and T_m_ value distribution for each target, respectively, which is beneficial for AMCA classification. Observed T_m_ values for *bla*_IMP_, *bla*_KPC_, *bla*_NDM_, *bla*_OXA-48_ and *bla*_VIM_ are 81.4, 89.5, 90.2, 83.8 and 87.9°C, respectively. Moreover, each primer set (in a multiplex environment) shows an excellent limit-of-detection (LOD) of 10 DNA copies/reaction. Corresponding standard curves, illustrating the C_q_ value as a function of the target concentration, yield an assay efficiency of 87.3, 103.5, 105.7, 98.7, 88.1%, respectively. PCR products were absent in all the negative controls.

#### 3.1.3 Experimental results in real-time dPCR

The 5-plex PCR assay was further validated in the dPCR platform with synthetic DNA templates at concentrations ranging from 10^1^ to 10^5^ DNA copies per panel, which were chosen such that we observe amplification events in both-single and bulk regions to capture kinetic information in both domains. Figure 2A shows end-point photographs (cycle 45) of panels at increasing amount of DNA. A total of 29,165 positive amplification reactions were performed. As shown in Figure 2B, a digital bulk standard curve for each target was build using the real-time dPCR instrument. As this microfluidic platform is capable of real-time data collection, quantification cycle values were used to generate the standard curves by plotting the C_q_ values against log[quantity] of a ten-fold serial dilution of each DNA target. It can be observed that there is a clear separation between the single-molecule (10^1^ to 10^2^ copies/panel) and the bulk regions (10^4^ to 10^5^ copies/panel) based on C_q_ value ranges, where 10^3^ copies/panel acts as a transition region across all the targets. In the none-saturated panels we can observed a digital pattern (number of ONs and OFFs) at the end of the reaction and the amount input molecules can be calculated using binomial and Poisson statistics (Quan et al., 2018), whereas in the saturated panels the amount input molecules can be quantified using the digital bulk standard curve (as in qPCR). Digital bulk standard curves yield an assay efficiency of 118.1, 98.7, 86.2, 100.8 and 90.2% efficiency for *bla*_IMP_, *bla*_KPC_, *bla*_NDM_, *bla*_OXA-48_ and *bla*_VIM_ assays, respectively. Table 2 reports the standard curve parameters for each assay, digital count and panel occupancy. Figures 3A and 3B, respectively, show the amplification and melting curves for the five carbapenem-resistant genes and the average characteristic sigmoidal shape for each target (black solid line) in real-time dPCR. Figure 3C represents the distribution of melting temperature, where the T_m_ range for each target is computed as: *bla*_IMP_ (81.3, 83.2°C), *bla*_KPC_ (89.0, 91.5°C), *bla*_NDM_ (90.0, 92.7°C), *bla*_OXA-48_ (83.7, 86.6°C) and *bla*_VIM_ (87.7, 90.8°C). After peak detection, negative reactions can be confirmed by identifying curves with no peak.

**Table 2.** Standard curve parameter in real-time digital PCR.

| Target | Slope | Constant | Rsqr<sup>a</sup> | Eff. (%)<sup>b</sup> | Single-molecule region |  | Transition region | Bulk region |  |
| --- | --- | --- | --- | --- | --- | --- | --- | --- | --- |
|  |  |  |  |  | 10<sup>1</sup> cp/pnl (occ.)<sup>c</sup> | 10<sup>2</sup> cp/pnl (occ.)<sup>c</sup> | 10<sup>3</sup> cp/pnl (occ.)<sup>c</sup> | 10<sup>4</sup> cp/pnl (occ.)<sup>c</sup> | 10<sup>5</sup> cp/pnl (occ.)<sup>c</sup> |
| *bla<sub>IMP</sub>* | -2.953 | 37.875 | 0.978 | 118.111 | 7 (0.9%) | 51 (6.6%) | 519 (67.4%) | 770 (100.0%) | 768 (99.7%) |
| *bla<sub>KPC</sub>* | -3.354 | 38.275 | 0.993 | 98.661 | 5 (0.6%) | 56 (7.3%) | 398 (51.7%) | 769 (99.9%) | 770 (100.0%) |
| *bla<sub>NDM</sub>* | -3.705 | 40.62 | 0.996 | 86.174 | 4 (0.5%) | 21 (2.7%) | 190 (24.7%) | 767 (99.6%) | 769 (99.9%) |
| *bla<sub>OXA-48</sub>* | -3.304 | 38.01 | 0.998 | 100.77 | 3 (0.4%) | 25 (3.2%) | 321 (41.7%) | 769 (99.9%) | 768 (99.7%) |
| *bla<sub>VIM</sub>* | -3.582 | 39.96 | 0.994 | 90.169 | 6 (0.8%) | 59 (7.7%) | 659 (85.6%) | 770 (100.0%) | 770 (100.0%) |

<sup>a</sup> R-squared<sup>b</sup> Efficiency (%)<sup>c</sup> copies/panel (% occupancy in digital PCR). The occupancy is calculated by counting the number of amplification reaction occurring per each panel and diving it by the total number of wells (N=770).

**Figure 2.**
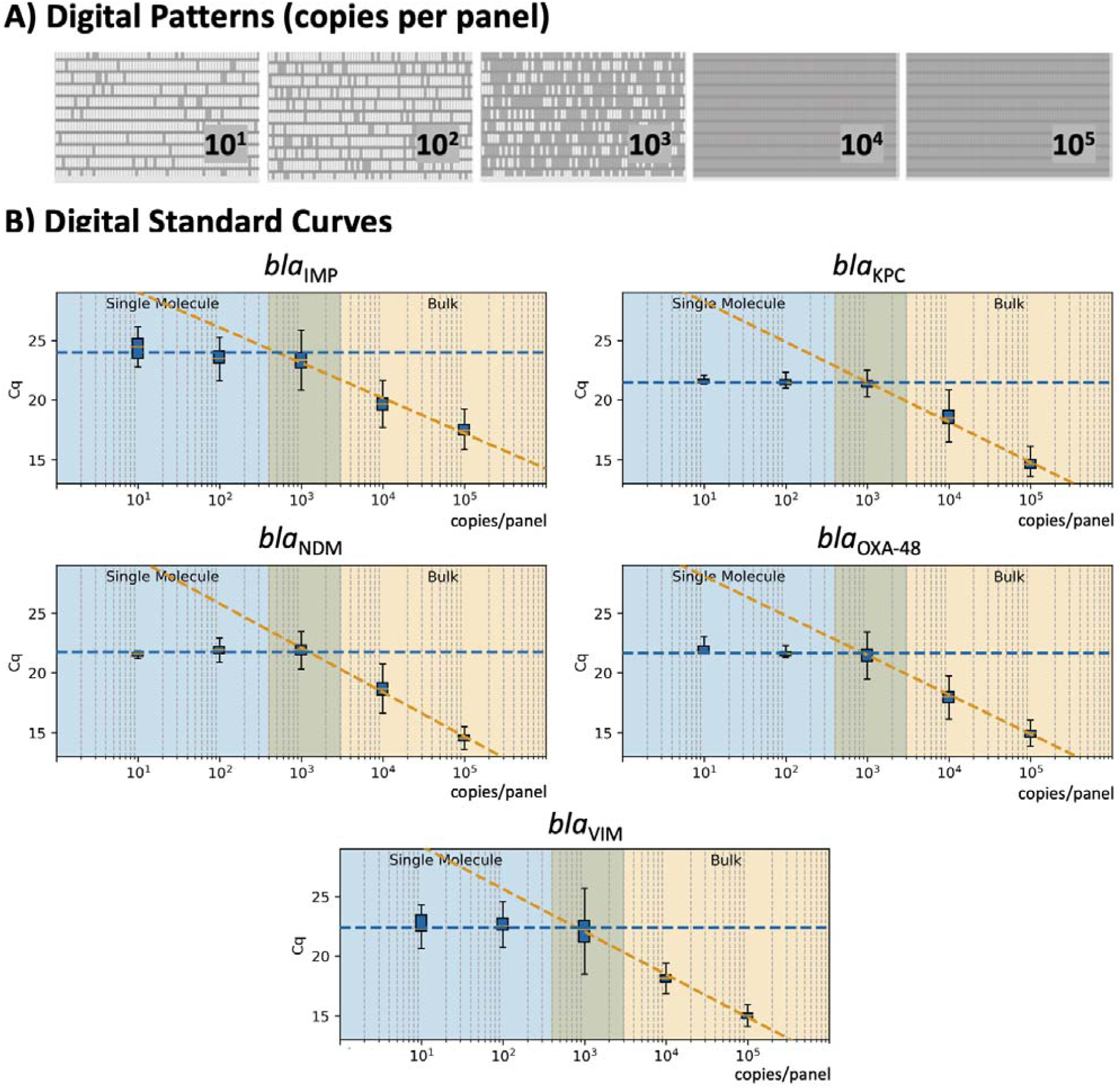
Standard Curve in real-time digital PCR. (A) Digital patterns for each microfluidic panel at increasing concentrations (770 reaction chambers per panel; 0.85 nL volume per chamber). (B) Standard curves correlating the C_q_ values with the concentration of each target; shaded blue area indicates the single-molecule region; shaded orange shows the bulk region; and the middle area displays the theoretical transition between the single-molecule and bulk.

**Figure 3.**
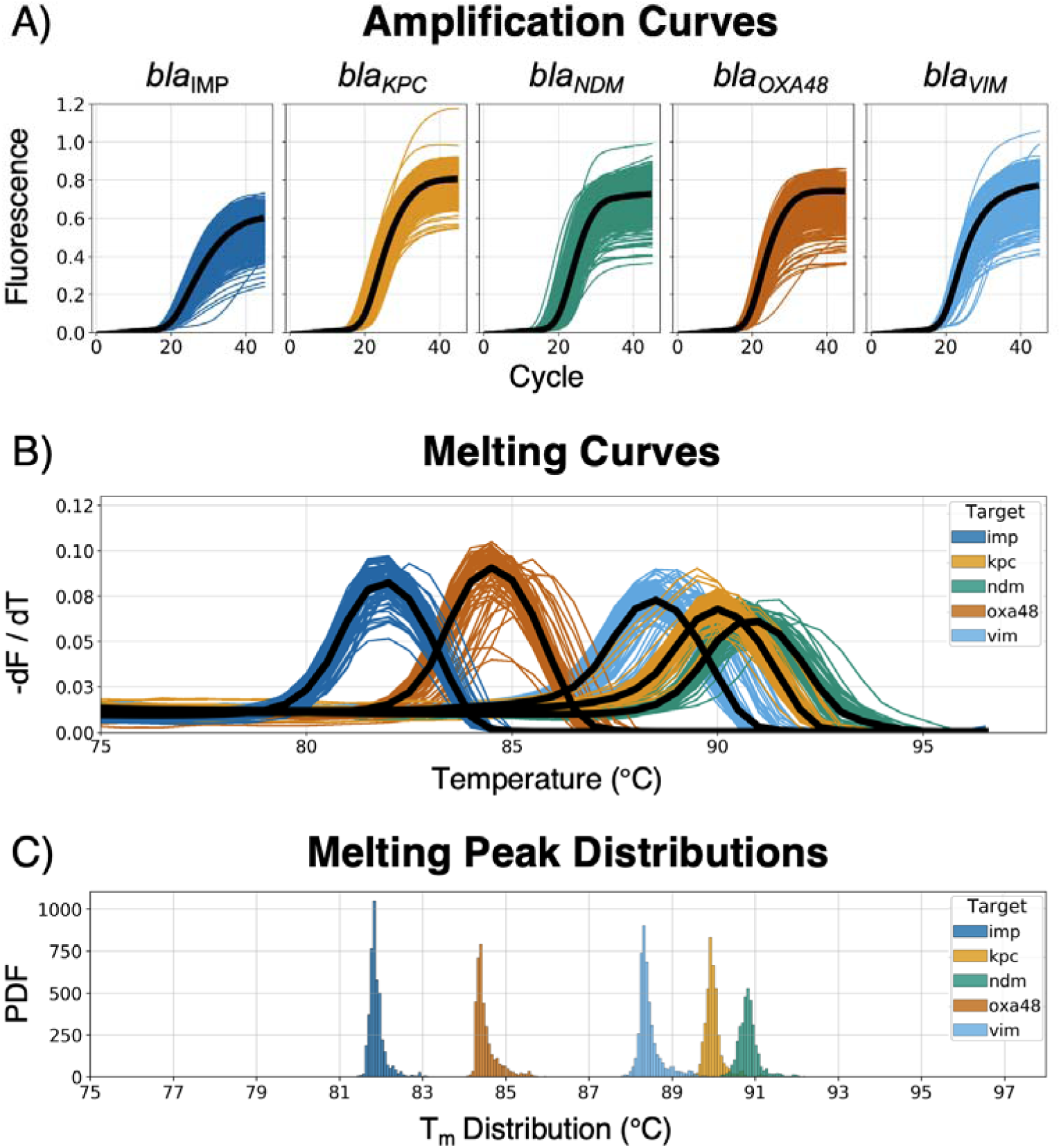
Real-time amplification and melting curves obtained from the dPCR instrument. (A) Raw amplification curves at different concentrations from synthetic DNA templates; the black line represents the average trend of the kinetic information based on each specific target-primer interaction. (B) Melting curves across the five different CPO; the black line represents the average trend of the thermodynamic information based on each specific target-primer interaction. (C) Melting peak (T_m_) distribution from the dPCR instrument, showing the probability density function (PDF) for each target.

### 3.2 Clinical isolates

As depicted in Table 3, the 253 pure bacterial strains were identified from MALDI-TOF MS as *Acinetobacter* spp. (n = 2), *Citrobacter* spp. (n = 16), *Enterobacter* spp. (n = 37), *Escherichia* spp. (n = 57), *Klebsiella* sp. (n = 133), *Proteu*s sp. (n = 1), *Pseudomonas* sp. (n = 5) and *Serratia* sp. (n = 2). Carbapenemase genes were determined as a single enzyme in 220 strains (*bla*_*IMP*_ = 45; *bla*_KPC_ = 9; *bla*_*NDM*_ = 74; *bla*_*OXA-48*_ = 84; *bla*_VIM_ = 8) and as a combination in one isolate (*bla*_*NDM*_ & *bla*_*OXA-48*_). Thirty-two isolates were confirmed as negative for the five carbapenemase genes. A more detailed description of each isolate, including bacterial species, date of sampling, specimen type, antibiotic resistance mechanisms and concentration (copies/µl of extracted DNA) can be found in Supplementary Table S1.

**Table 3.** Clinical Enterobacteriaceae isolates used in this study.

| Species (MALDI-TOF MS) | Carbapenemase gene | Number of isolates |
| --- | --- | --- |
| *Citrobacter* spp. | *bla<sub>IMP</sub>* | 1 |
|  | *bla<sub>KPC</sub>* | 2 |
|  | *bla<sub>NDM</sub>* | 1 |
|  | *bla<sub>OXA-48</sub>* | 10 |
|  | *bla<sub>VIM</sub>* | 1 |
| *Enterobacter* spp. | *bla<sub>IMP</sub>* | 20 |
|  | *bla<sub>NDM</sub>* | 7 |
|  | *bla<sub>OXA-48</sub>* | 2 |
|  | *bla<sub>VIM</sub>* | 2 |
| *Escherichia* spp. | *bla<sub>IMP</sub>* | 7 |
|  | *bla<sub>NDM</sub>* | 14 |
|  | *bla<sub>NDM</sub> & bla<sub>OXA-48</sub>* | 1 |
|  | *bla<sub>OXA-48</sub>* | 26 |
| *Klebsiella pneumoniae* | *bla<sub>IMP</sub>* | 15 |
|  | *bla<sub>KPC</sub>* | 6 |
|  | *bla<sub>NDM</sub>* | 51 |
|  | *bla<sub>OXA-48</sub>* | 45 |
|  | *bla<sub>VIM</sub>* | 3 |
| *Proteus mirabilis* | *bla<sub>NDM</sub>* | 1 |
| *Pseudomonas aeruginosa* | *bla<sub>IMP</sub>* | 2 |
|  | *bla<sub>VIM</sub>* | 2 |
| *Serratia marcescens* | *bla<sub>KPC</sub>* | 1 |
|  | *bla<sub>OXA-48</sub>* | 1 |
| Multiple species* | negative | 32 |

\*CPO-negative species: *Acinetobacter baumannii*, *Citrobacter freundii*, *Enterobacter* spp., *Escherichia coli*, *Klebsiella pneumoniae* and *Pseudomonas aeruginosa*.

### 3.3 The AMCA model: training and cross-validation

Our study aims to validate the performance of the AMCA method for detection of carbapenem-resistant genes in clinical isolates compared with the MCA approach. To train both models, a total of 99,860 amplification events were generated using synthetic DNA templates, of which 29,165 were positive: *bla*_IMP_ (N=4,941), *bla*_KPC_ (N=5,940), *bla*_NDM_ (N=5,870), *bla*_OXA-48_ (N=4,333) and *bla*_VIM_ (N=8,081). Observed overall classification performance of training dataset for the MCA and AMCA methods was 94.9 ± 21.99% and 99.2% ± 8.86%, respectively. Supplementary Figure S8 shows the confusion matrices comparing the true and predicted targets for both methods. It can be observed that the *bla*_NDM_ and *bla*_KPC_ targets are misclassified by the MCA methods, whereas the AMCA considerably improves the prediction of both targets: from 804 to 52 amplification events for *bla*_NDM,_ and from 511 to 46 for *bla*_KPC_. No other target was misclassified more than 1.26% for either method.

### 3.4 The AMCA model: clinical validation

A total of 253 clinical isolates, including 221 positives, and 224,840 amplification events (of which 160,041 positives) were used for the clinical validation. Compare to results obtained with the Xpert Carba-R Cepheid and Resist-3 O.K.N assays, the overall observed accuracy for MCA was 91.7% (CI 87.59 to 94.79%) and 99.6% (CI 97.82% to 99.99%) for AMCA, which represent a 7.9% increase (p-value < 0.01) (Supplementary Figure S9). A total of 21 clinical isolates were misclassified for the MCA method and considered false positives (FP) as shown in Table 4, whereas the AMCA reduced the number of misclassified samples to 1 (Table 5). All the false positive samples were identified as double infection because of the overlapping distribution in the T_m_, as shown in Figure 3. Performance improvement in the AMCA method is due to the addition of real-time amplification data, contrary to the MCA approach that only takes into account the melting curve distribution. Further details on AMCA coefficient contributions (i.e., ACA and MCA weights) are shown in Supplementary Figure S10. Moreover, 32 bacterial isolates not carrying the five carbapenemase genes were used to evaluate the assay specificity. The 5-plex PCR assay showed negative results in the absence of the specific target.

**Table 4.**
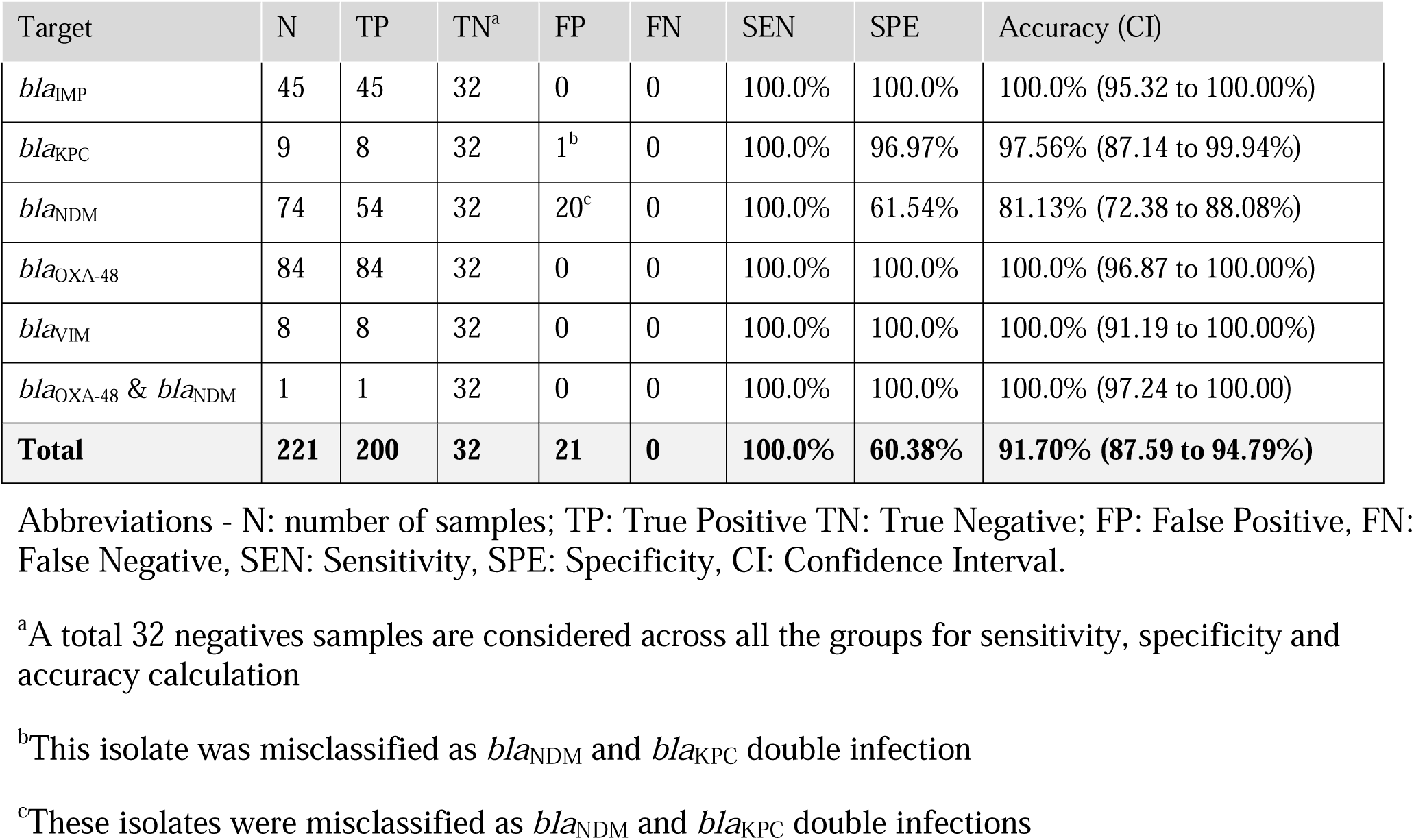
Classification of clinical isolates when using the ML-based MCA method

| Target | N | TP | TN<sup>a</sup> | FP | FN | SEN | SPE | Accuracy (CI) |
| --- | --- | --- | --- | --- | --- | --- | --- | --- |
| *bla*<sub>IMP</sub> | 45 | 45 | 32 | 0 | 0 | 100.0% | 100.0% | 100.0% (95.32 to 100.00%) |
| *bla*<sub>KPC</sub> | 9 | 8 | 32 | 1<sup>b</sup> | 0 | 100.0% | 96.97% | 97.56% (87.14 to 99.94%) |
| *bla*<sub>NDM</sub> | 74 | 54 | 32 | 20<sup>c</sup> | 0 | 100.0% | 61.54% | 81.13% (72.38 to 88.08%) |
| *bla*<sub>OXA-48</sub> | 84 | 84 | 32 | 0 | 0 | 100.0% | 100.0% | 100.0% (96.87 to 100.00%) |
| *bla*<sub>VIM</sub> | 8 | 8 | 32 | 0 | 0 | 100.0% | 100.0% | 100.0% (91.19 to 100.00%) |
| *bla*<sub>OXA-48</sub> & *bla*<sub>NDM</sub> | 1 | 1 | 32 | 0 | 0 | 100.0% | 100.0% | 100.0% (97.24 to 100.00) |
| **Total** | **221** | **200** | **32** | **21** | **0** | **100.0%** | **60.38%** | **91.70% (87.59 to 94.79%)** |

Abbreviations - N: number of samples; TP: True Positive TN: True Negative; FP: False Positive, FN: False Negative, SEN: Sensitivity, SPE: Specificity, CI: Confidence Interval.
<sup>a</sup>A total 32 negatives samples are considered across all the groups for sensitivity, specificity and accuracy calculation
<sup>b</sup>This isolate was misclassified as *bla*<sub>NDM</sub> and *bla*<sub>KPC</sub> double infection
<sup>c</sup>These isolates were misclassified as *bla*<sub>NDM</sub> and *bla*<sub>KPC</sub> double infections

**Table 5.**
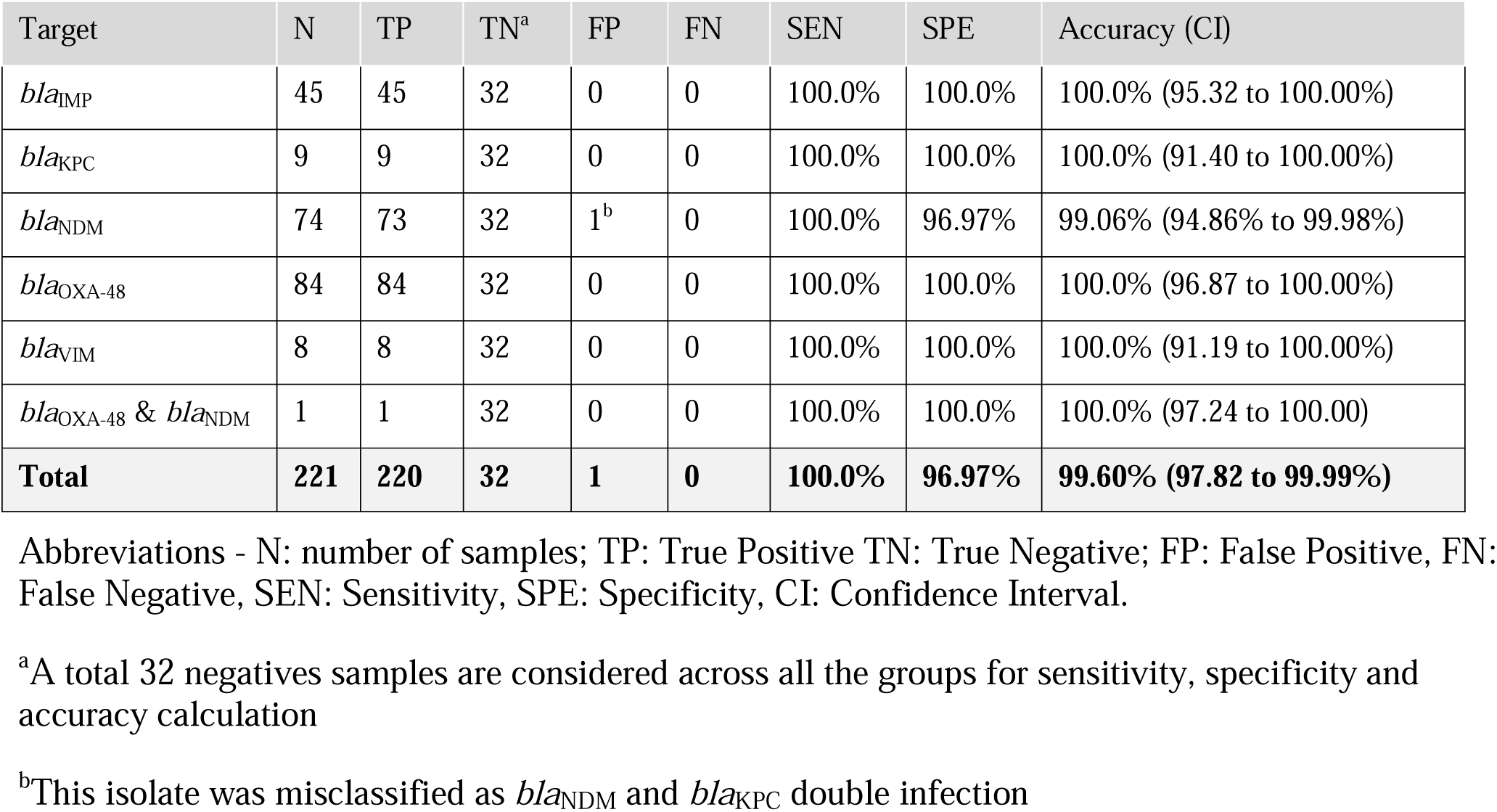
Classification of clinical isolates based on ML-based AMCA method

## 4 Discussion

In the last decade, novel pandemic outbreaks and the continued threats of emerging multi-drug resistant microorganisms have significantly increased the demand for molecular tests, in particular PCR-based methods (Nishizawa and Suzuki, 2014; Vasala et al., 2020). To respond to this need, the AMCA technology has been designed to increase the throughput of real-time molecular platforms. Seamlessly integrated with conventional diagnostic workflows, this machine learning based approach can enhance multiplexing capabilities of traditional qPCR and state-of-the art dPCR instruments, increasing the number of nucleic acid targets that can be identified in a single fluorescent channel without hardware modifications. Individual primer sets produce amplification products at a sequence-specific amplification rate and efficiency, which generate unique amplification and melting curves for different target concentrations. Such curves can be capture as time-series data by real-time instruments, feed into machine learning models and used to identify multidimensional patterns (or signatures) specific to each primer set. Therefore, enabling the identification of multiple DNA targets per fluorescent channel using only real-time data (i.e., data-driven multiplexing). In this paper, we performed a clinical validation on diagnostic accuracy of the AMCA methodology by targeting the “big five” carbapenem-resistant genes (*bla*_VIM_, *bla*_OXA-48_, *bla*_NDM_, *bla*_IMP_ and *bla*_KPC_) in multiplex PCR. A 5-plex PCR assay was developed and characterised in both real-time qPCR and dPCR instruments, and the AMCA performance investigated through the identification of 253 clinical isolates from patients’ samples. The MCA was used as a reference method to compare results.

We successfully show a 99.2% accuracy for identifying the five carbapenem-resistant genes in the clinical isolates. The AMCA method was shown to enhance the classification performance by 7.9% compared to MCA. The AMCA takes advantage of the volume of raw data extracted from amplification and melting curves, whereas the MCA only considers melting curves. It is interesting to observe that the overlapping melting curve distribution in Figure 3B (e.g. *bla*_NDM_ and *bla*_KPC_) represents a misclassification of 1303 reactions (509 *bla*_KPC_ as *bla*_NDM_, and 804 *bla*_NDM_ as *bla*_KPC_) and 21 clinical isolates (20 *bla*_NDM_ and 1 *bla*_KPC_ as coinfections) when using the MCA, but it only represents a misclassification of 99 reactions and 1 clinical isolates for the AMCA method. As described in previous publications (Moniri et al., 2020a), these results support the hypothesis that the underlaying biological factors driving these methods for target identification are fundamentally different. As observed in Supplementary Figure S10, machine learning methods can be used to exploit the distinctive information contained on the amplification and melting curves by weighting the predictions from the ACA and MCA to optimally combine them and maximize the AMCA performance.

Although dPCR is not likely to replace all qPCR assays in the clinical laboratory due to associated instrument costs and greater complexity, it has several specific advantages over qPCR. The vast number of partitions reduce the likelihood of coamplification and inhibitors in a single reaction, facilitating accurate detection of multiple analytes; and the large amount of data enables the use of advance machine learning algorithms to detect subtle kinetic and thermodynamic differences encoded in the real-time amplification data. On the other hand, real-time dPCR platforms enable the use of digital bulk standards and offer a valuable solution for absolute quantification of clinical isolates (equivalently to conventional qPCR standards) even when the panels are saturated, expanding the dynamic range of quantification of the microfluidic chips and eliminating the need of testing the samples at multiple dilutions to ensure that at least one of them falls within the conventional dPCR range (i.e. panels at occupancy <85%). As shown in Figure 2, it is possible to create a standard curve in real-time dPCR by extracting C_q_ values as a function of the target concentration because there is a clear separation between the single-molecule and the bulk regions. We envision that coupling real-time dPCR instruments with data-driven multiplexing will expand the use of these platforms in clinical microbiology laboratories.

The results presented in this study represent a step forward in the use of PCR-based data-driven diagnostics for clinical applications. However, there are several aspects that need to be further investigated. Firstly, in this paper we evaluated the performance of AMCA method in clinical isolates using pure bacterial cultures, therefore a follow-up study needs to be conducted to evaluate the performance of the method directly from clinical samples. Secondly, it is important to identify co-presence of infections for patient treatment, however in this paper we address only one sample with a double infection; a larger study will be required to test the effectiveness of the AMCA in double pathogen identification. Depending on the sample concentration, this might not limit multiplexing capabilities in dPCR, but it could represent a challenge when qPCR instruments are used.

This work suggests that the AMCA approach provides a versatile solution for the accurate detection of AMR genes, representing a cost-effective interaction as it does not require hardware modifications. This study highlights the importance of integrating artificial intelligence for diagnosis and how effectively it increases result reliability of state-of-the-art dPCR instruments. Moreover, the AMCA methodology has the potential for further application in point-of-care devices and isothermal chemistries, as a solution to leverage identification accuracy and enable faster detection of multiple pathogens.

## 5 Data availability statement

The python code and datasets generated for this study can be found in GitHub repository at: https://github.com/LMigliet/pyAMCA_5plex.

## Data Availability

All data referred in the manuscript is available and/or provided in the supplementary information.

## 7 Acknowledgments

We thank the staff of the diagnostic microbiology laboratory of North West London Pathology for isolate collection and storage.

## 8 Supplementary material

The Supplementary Material for this article can be found online

## References

1. Athamanolap, P., Parekh, V., Fraley, S. I., Agarwal, V., Shin, D. J., Jacobs, M. A., et al. (2014). Trainable high resolution melt curve machine learning classifier for large-scale reliable genotyping of sequence variants. PLoS One 9. doi:10.1371/journal.pone.0109094.

2. Beinhauerova, M., Babak, V., Bertasi, B., Boniotti, M. B., and Kralik, P. (2020). Utilization of Digital PCR in Quantity Verification of Plasmid Standards Used in Quantitative PCR. Front. Mol. Biosci. 7, 1–13. doi:10.3389/fmolb.2020.00155.

3. Bleumin, D., Cohen, M. J., Moranne, O., Esnault, V. L. M., Benenson, S., Paltiel, O., et al. (2012). Carbapenem-resistant Klebsiella pneumoniae is associated with poor outcome in hemodialysis patients. J. Infect. 65, 318–325. doi:10.1016/j.jinf.2012.06.005.

4. Bush, K., and Fisher, J. F. (2011). Epidemiological Expansion, Structural Studies, and Clinical Challenges of New β-Lactamases from Gram-Negative Bacteria. Annu. Rev. Microbiol. 65, 455–478. doi:10.1146/annurev-micro-090110-102911.

5. Charani, E., McKee, M., Ahmad, R., Balasegaram, M., Bonaconsa, C., Merrett, G. B., et al. (2021). Optimising antimicrobial use in humans – review of current evidence and an interdisciplinary consensus on key priorities for research. Lancet Reg. Heal. - Eur. 7, 100161. doi:10.1016/j.lanepe.2021.100161.

6. Codjoe, F., and Donkor, E. (2017). Carbapenem Resistance: A Review. Med. Sci. 6, 1. doi:10.3390/medsci6010001.

7. Collins, G. S., and Moons, K. G. M. (2019). Reporting of artificial intelligence prediction models. Lancet 393, 1577–1579. doi:10.1016/S0140-6736(19)30037-6.

8. Cunningham, P., and Delany, S. J. (2020). k-Nearest Neighbour Classifiers: 2nd Edition (with Python examples). 1–22. doi:10.1145/3459665.

9. Edgar, R. C. (2004). MUSCLE: Multiple sequence alignment with high accuracy and high throughput. Nucleic Acids Res. 32, 1792–1797. doi:10.1093/nar/gkh340.

10. Eliopoulos, G. M., Schwaber, M. J., and Carmeli, Y. (2014). An ongoing national intervention to contain the spread of carbapenem-resistant enterobacteriaceae. Clin. Infect. Dis. 58, 697–703. doi:10.1093/cid/cit795.

11. Huggett, J. F., Cowen, S., and Foy, C. A. (2015). Considerations for digital PCR as an accurate molecular diagnostic tool. Clin. Chem. 61, 79–88. doi:10.1373/clinchem.2014.221366.

12. Johnning, A., Karami, N., Tång Hallbäck, E., Müller, V., Nyberg, L., Buongermino Pereira, M., et al. (2018). The resistomes of six carbapenem-resistant pathogens - a critical genotype-phenotype analysis. Microb. genomics 4. doi:10.1099/mgen.0.000233.

13. Lim, Y. J., Park, H. Y., Lee, J. Y., Kwak, S. H., Kim, M. N., Sung, H., et al. (2018). Clearance of carbapenemase-producing Enterobacteriaceae (CPE) carriage: a comparative study of NDM-1 and KPC CPE. Clin. Microbiol. Infect. 24, 1104.e5-1104.e8. doi:10.1016/j.cmi.2018.05.013.

14. Matsumura, Y., and Pitout, J. D. (2016). Recent advances in the laboratory detection of carbapenemase-producing Enterobacteriaceae. Expert Rev. Mol. Diagn. 16, 783–794. doi:10.1586/14737159.2016.1172964.

15. Maurer, F. P., Castelberg, C., Quiblier, C., Bloemberg, G. V., and Hombach, M. (2015). Evaluation of carbapenemase screening and confirmation tests with Enterobacteriaceae and development of a practical diagnostic algorithm. J. Clin. Microbiol. 53, 95–104. doi:10.1128/JCM.01692-14.

16. Mercaldo ND, Lau KF, Z. X. (2007). Confidence intervals for predictive values with an emphasis to. Stat Med 26, 2170–2183. doi:10.1002/sim.2677.

17. Moloney, E., Lee, K. W., Craig, D., Allen, A. J., Graziadio, S., Power, M., et al. (2019). A PCR-based diagnostic testing strategy to identify carbapenemase-producing Enterobacteriaceae carriers upon admission to UK hospitals: early economic modelling to assess costs and consequences. Diagnostic Progn. Res. 3, 1–9. doi:10.1186/s41512-019-0053-x.

18. Moniri, A., Miglietta, L., Holmes, A., Georgiou, P., and Rodriguez-Manzano, J. (2020a). High-Level Multiplexing in Digital PCR with Intercalating Dyes by Coupling Real-Time Kinetics and Melting Curve Analysis. Anal. Chem. 92, 14181–14188. doi:10.1021/acs.analchem.0c03298.

19. Moniri, A., Miglietta, L., Malpartida-Cardenas, K., Pennisi, I., Cacho-Soblechero, M., Moser, N., et al. (2020b). Amplification Curve Analysis: Data-Driven Multiplexing Using Real-Time Digital PCR. Anal. Chem. 92, 13134–13143. doi:10.1021/acs.analchem.0c02253.

20. Moniri, A., Rodriguez-Manzano, J., Malpartida-Cardenas, K., Yu, L. S., Didelot, X., Holmes, A., et al. (2019). Framework for DNA Quantification and Outlier Detection Using Multidimensional Standard Curves. Anal. Chem. 91, 7426–7434. doi:10.1021/acs.analchem.9b01466.

21. multiple-primer-analyzer @ www.thermofisher.com Available at: https://www.thermofisher.com/uk/en/home/brands/thermo-scientific/molecular-biology/molecular-biology-learning-center/molecular-biology-resource-library/thermo-scientific-web-tools/multiple-primer-analyzer.html.

22. Neuner, E. A., Yeh, J. Y., Hall, G. S., Sekeres, J., Endimiani, A., Bonomo, R. A., et al. (2011). Treatment and outcomes in carbapenem-resistant Klebsiella pneumoniae bloodstream infections. Diagn. Microbiol. Infect. Dis. 69, 357–362. doi:10.1016/j.diagmicrobio.2010.10.013.

23. Nishizawa, T., and Suzuki, H. (2014). Mechanisms of Helicobacter pylori antibiotic resistance and molecular testing. Front. Mol. Biosci. 1, 1–7. doi:10.3389/fmolb.2014.00019.

24. Otter, J. A., Burgess, P., Davies, F., Mookerjee, S., Singleton, J., Gilchrist, M., et al. (2017a). Counting the cost of an outbreak of carbapenemase-producing Enterobacteriaceae: an economic evaluation from a hospital perspective. Clin. Microbiol. Infect. 23, 188–196. doi:10.1016/j.cmi.2016.10.005.

25. Otter, J. A., Doumith, M., Davies, F., Mookerjee, S., Dyakova, E., Gilchrist, M., et al. (2017b). Emergence and clonal spread of colistin resistance due to multiple mutational mechanisms in carbapenemase-producing Klebsiella pneumoniae in London. Sci. Rep. 7, 1–8. doi:10.1038/s41598-017-12637-4.

26. Quan, P. L., Sauzade, M., and Brouzes, E. (2018). DPCR: A technology review. Sensors (Switzerland) 18. doi:10.3390/s18041271.

27. Reta, D. H., Tessema, T. S., Ashenef, A. S., Desta, A. F., Labisso, W. L., Gizaw, S. T., et al. (2020). Molecular and Immunological Diagnostic Techniques of Medical Viruses. Int. J. Microbiol. 2020. doi:10.1155/2020/8832728.

28. Rodriguez-Manzano, J., Moniri, A., Malpartida-Cardenas, K., Dronavalli, J., Davies, F., Holmes, A., et al. (2019). Simultaneous Single-Channel Multiplexing and Quantification of Carbapenem-Resistant Genes Using Multidimensional Standard Curves. Anal. Chem. 91, 2013–2020. doi:10.1021/acs.analchem.8b04412.

29. Rodriguez-Manzano, J., Moser, N., Malpartida-Cardenas, K., Moniri, A., Fisarova, L., Pennisi, I., et al. (2020). Rapid Detection of Mobilized Colistin Resistance using a Nucleic Acid Based Lab-on-a-Chip Diagnostic System. Sci. Rep. 10, 1–9. doi:10.1038/s41598-020-64612-1.

30. Tzouvelekis, L. S., Markogiannakis, A., Psichogiou, M., Tassios, P. T., and Daikos, G. L. (2012). Carbapenemases in Klebsiella pneumoniae and other Enterobacteriaceae: An evolving crisis of global dimensions. Clin. Microbiol. Rev. 25, 682–707. doi:10.1128/CMR.05035-11.

31. Vasala, A., Hytönen, V. P., and Laitinen, O. H. (2020). Modern Tools for Rapid Diagnostics of Antimicrobial Resistance. Front. Cell. Infect. Microbiol. 10. doi:10.3389/fcimb.2020.00308.

