## Supplementary material for "Coupling machine learning and high throughput multiplex digital PCR enables accurate detection of carbapenem-resistant genes in clinical isolates"

### 1 Supplementary Data

#### 1.1 Supplementary Data S1

*Experimental condition for Real-Time PCR*: Each amplification reaction was performed in 10 μL of final volume with 5 μL of 2× SsoFast EvaGreen Supermix with Low ROX (BioRad, UK), 3 μL of PCR-grade water, 1 μL of 10× multiplex PCR primer mixture containing the five primer sets (5μM of each primer), and 1μL of different concentrations of synthetic DNA, clinical sample or controls. The reaction consisted of 10 min at 95 °C, followed by 45 cycles at 95 °C for 20 s, 67 °C for 45 s, and 72 °C for 30 s. Melting curve analysis was performed with one cycle at 65 °C for 60 s and reading from 65 to 97 °C with an increment of 0.2 °C. The PCR machine used in this study was the Light Cycler 96 real-time PCR system (Roche Diagnostics, Germany).

#### 1.2 Supplementary Data S2

*AMCA coefficient equation*: As the colourmap in Supplementary Figure S10 shows, the Amplification Curve Analysis (ACA) and Melting Curve Analysis (MCA) coefficients contribute differently to the target classification of the Amplification and Melting Curve Analysis (AMCA) model. More specifically, because the AMCA is a supervised linear model, the coefficients can be investigated to understand how it weighs the predictions from ACA and MCA. The output of AMCA is defined by:

$$y=\hat{W}_{ACA} y_{ACA}+ \hat{W}_{MCA} y_{MCA}$$

where $y_{ACA}\epsilon\mathbb{R}^{5}$ and $y_{MCA}\epsilon\mathbb{R}^{5}$ are the probabilities outputted from the ACA and MCA models, respectively, and $\hat{W}_{ACA} \epsilon\mathbb{R}^{5X5}$ and $\hat{W}_{MCA} \epsilon\mathbb{R}^{5X5}$ are the model coefficients relating to ACA and MCA, respectively.

### 2 Supplementary Figures and Tables

#### 2.1 Supplementary Figures

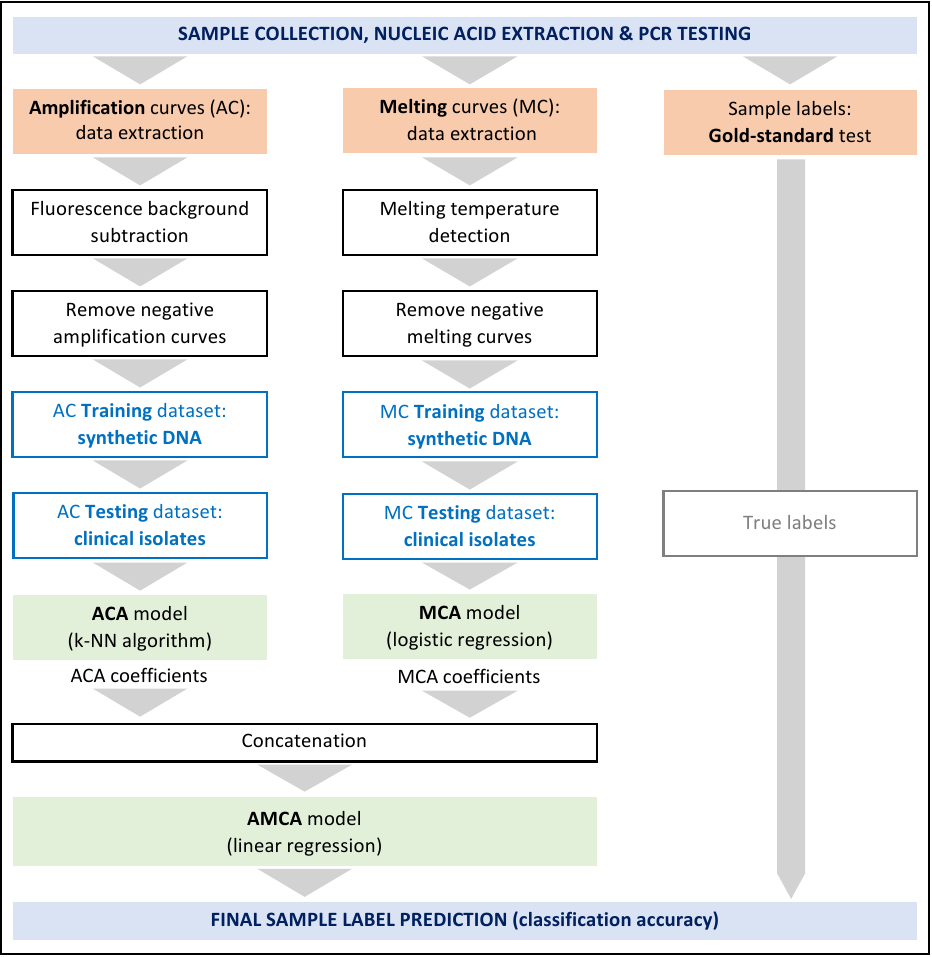

**Supplementary Figure S1**. Flowchart to visualize the data processing workflow for the AMCA method. The true label (determined by a gold standard PCR method such as Cepheid Xpert Carba-R assay) are only required as comparison for the predicated labels from the AMCA algorithm. The training data set used for the ACA and MCA model is generated using synthetic DNA with known labels and concentrations, whereas the testing dataset is generated from clinical isolates dataset using digital real-time PCR. The output of ACA and MCA models are probabilities for each target and their coefficients are concatenated as input of the final AMCA model.

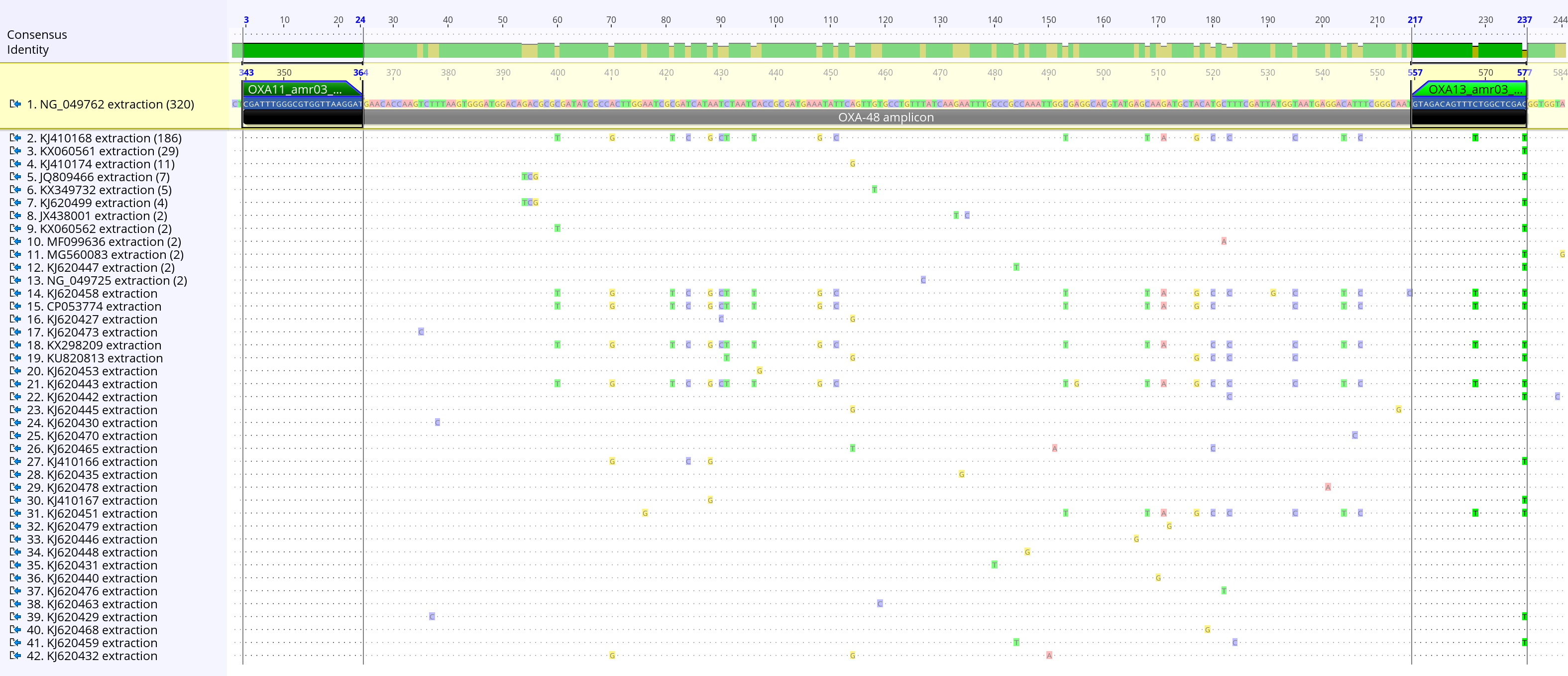

**Supplementary Figure S2**. *Inclusivity alignment of* *bla*_OXA-48_. Sequences retrieved from nr/nt NCBI database (n = 603) with a coverage of 100% for each primer binding region. The alignment shows only unique sequences that differ from the reference NG_049762 in the amplification region. The sequences are ordered from the largest number of unique sequences to the lowest as shown in bracket (after the NCBI accession number) on the left side of the alignment.

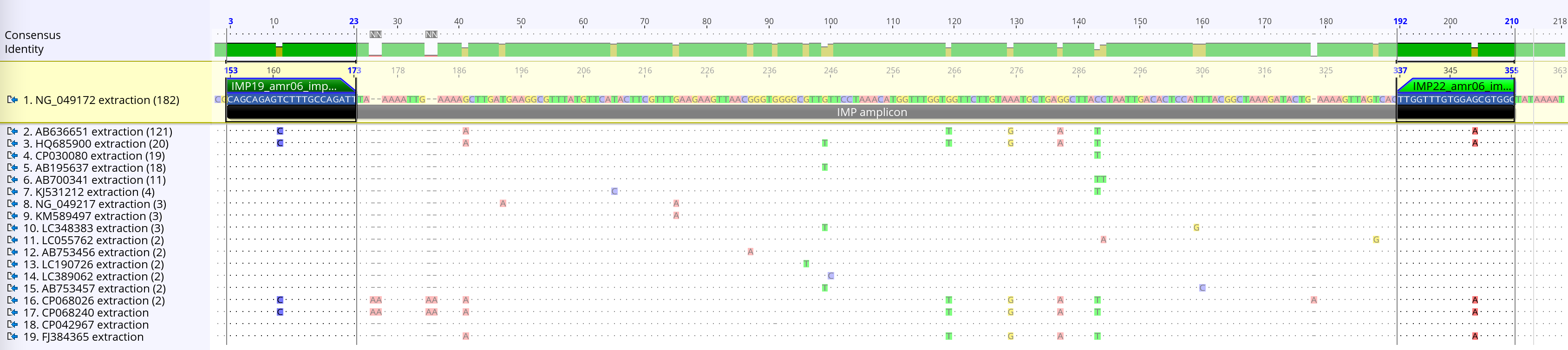

**Supplementary Figure S3**. *Inclusivity alignment of bla*_IMP_ (including bla_IMP-1_ and bla_IMP-4_ groups). Sequences retrieved from nr/nt NCBI database (n = 400) with a coverage of 100% for each primer binding region. The alignment shows only unique sequences that differ from the reference NG_049172 in the amplification region. The sequences are ordered from the largest number of unique sequences to the lowest as shown in bracket (after the NCBI accession number) on the left side of the alignment.

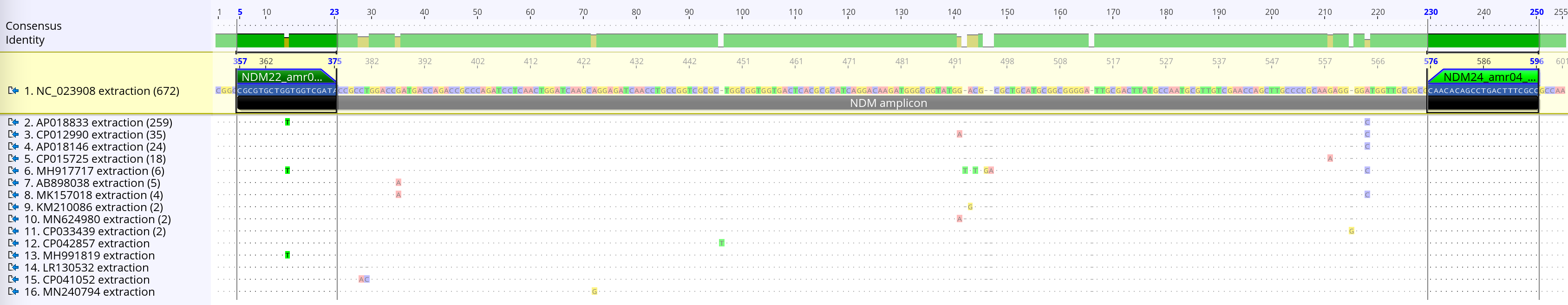

**Supplementary Figure S4**. *Inclusivity alignment of bla*_NDM_. Sequences retrieved from nr/nt NCBI database (n = 1035) with a coverage of 100% for each primer binding region. The alignment shows only unique sequences that differ from the reference NC_023908 in the amplification region. The sequences are ordered from the largest number of unique sequences to the lowest as shown in bracket (after the NCBI accession number) on the left side of the alignment.

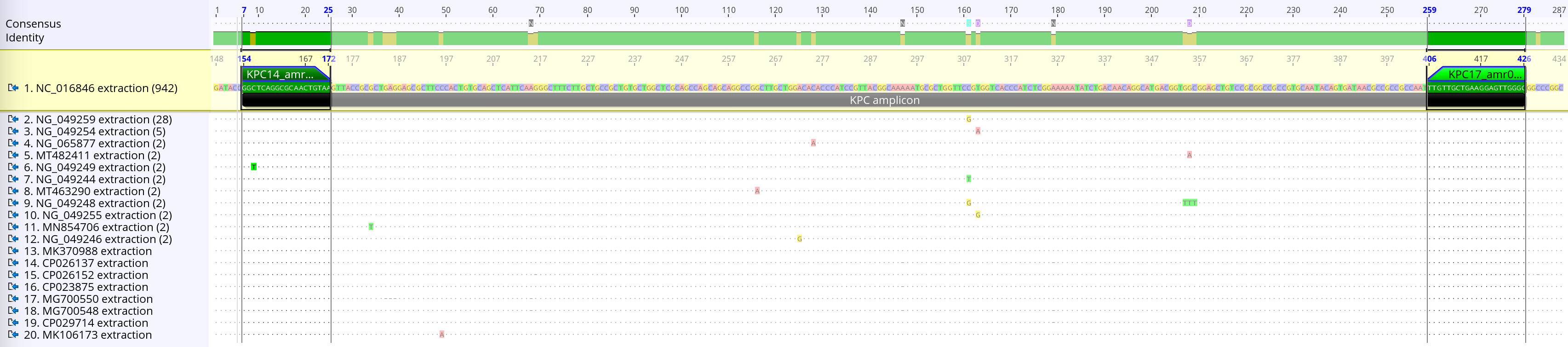

**Supplementary Figure S5**. *Inclusivity alignment of bla*_KPC_. Sequences retrieved from nr/nt NCBI database (n = 1001) with a coverage of 99.9% for each primer binding region. The alignment shows only unique sequences that differ from the reference NC_016846 in the amplification region. The sequences are ordered from the largest number of unique sequences to the lowest as shown in bracket (after the NCBI accession number) on the left side of the alignment.

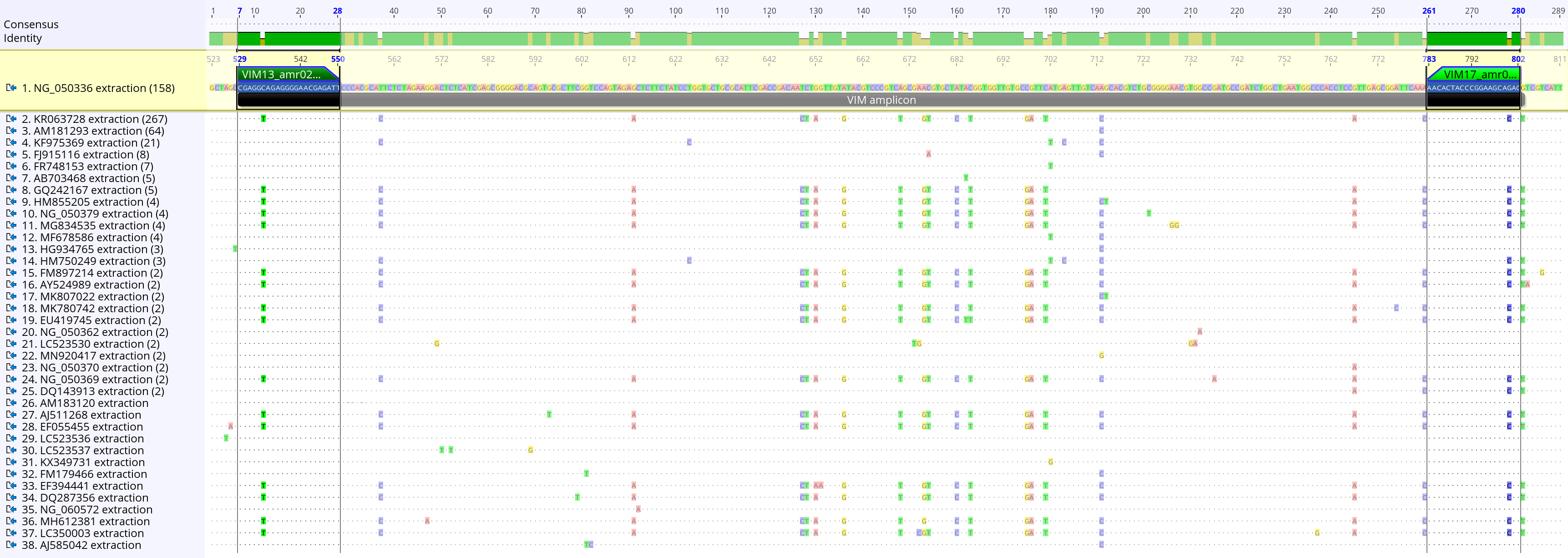

**Supplementary Figure S6**. *Inclusivity alignment of bla*_VIM_. Sequences retrieved from nr/nt NCBI database (n = 593) with a coverage of 100% for each primer binding region. The alignment shows only unique sequences that differ from the reference NG_050336 in the amplification region. The sequences are ordered from the largest number of unique sequences to the lowest as shown in bracket (after the NCBI accession number) on the left side of the alignment.

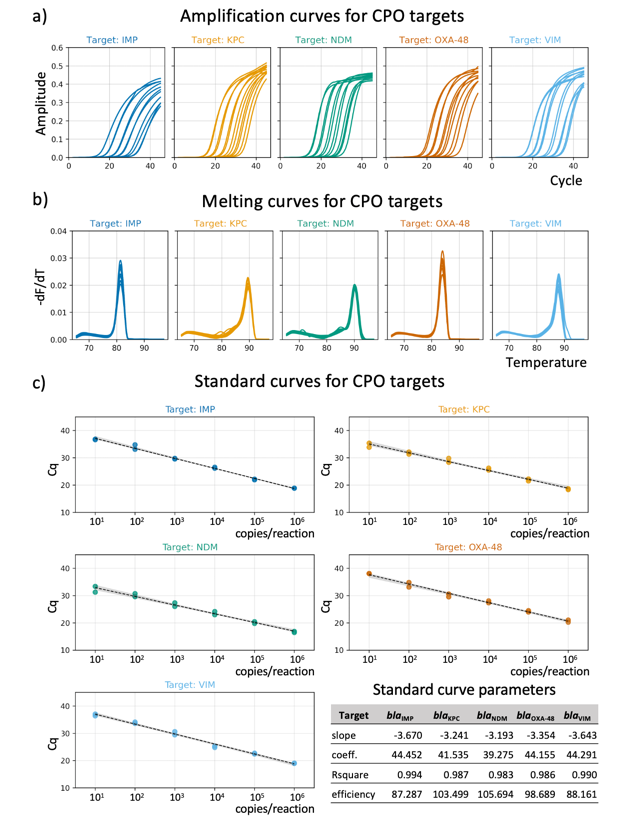

**Supplementary Figure S7**. Analysis of real-time amplification and melting curves from qPCR instruments. (a) Raw real-time amplification curves. (b) Raw melting curve for each target. (c) Standard curves for each target using our new 5-plex PCR assay. (Bottom Panel) Table with relevant meta data for each standard curve.

**Supplementary Figure S8**. *Performance of the MCA and AMCA methods for multiplexing the five carbapenemase gene targets in the training dataset using synthetic DNA templates*. (left) Confusion matrix illustrating the predictions from Melting Curve Analysis (MCA) algorithm. (right) Confusion matrix illustrating the predictions from Amplification and Melting Curve Analysis (AMCA) algorithm. Values in the matrices indicate the number of positive amplification events (N=29,165) with diagonal entries corresponding to correct predictions.

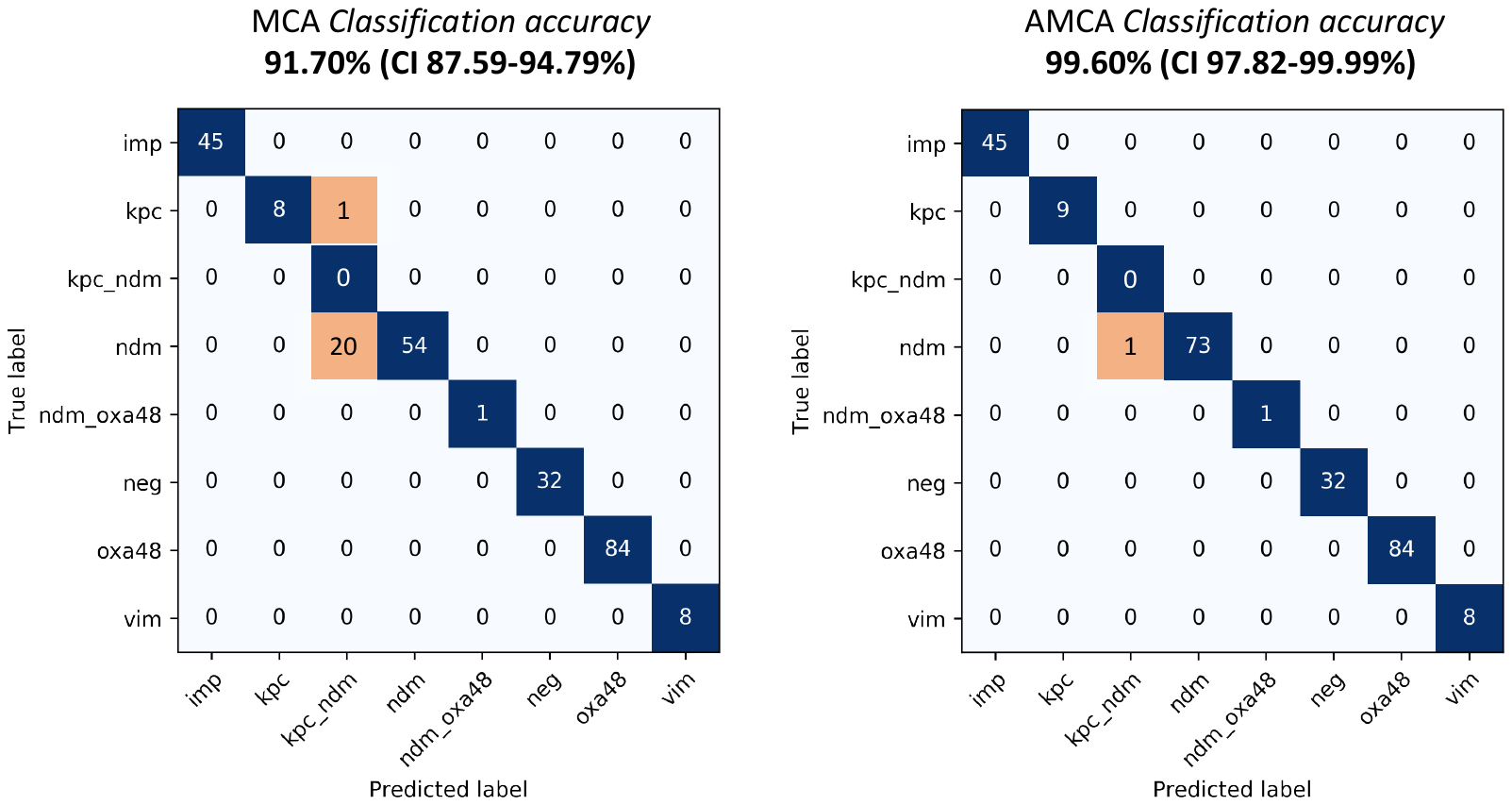

**Supplementary Figure S9**. *Performance of MCA and AMCA methods for multiplexing the five carbapenemase gene targets in clinical isolates*. (left) Confusion matrix illustrating the predictions from Melting Curve Analysis (MCA) algorithm. (right) Confusion matrix illustrating the predictions from Amplification and Melting Curve Analysis (AMCA) algorithm. Values indicate the number of clinical isolates (N=253) with diagonal entries corresponding to correct predictions.

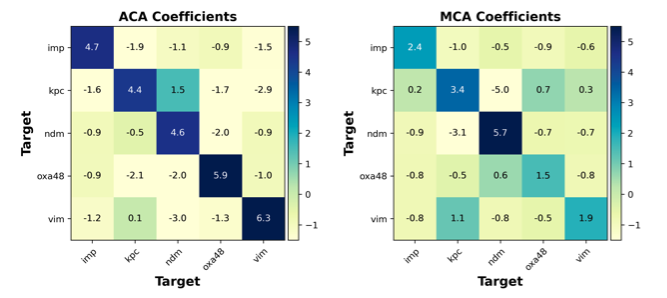

**Supplementary Figure S10.** *The coefficients of the AMCA model*. The values in the confusion matrices, ranging from -5 to 6.3, indicate the predictions weights from the ACA and MCA methods, respectively. Darker colours indicate more positive weighting. For example, as it can be observed, the AMCA weighs the prediction from ACA more heavily for *bla*_IMP_ target (4.7 in the ACA model compared to the 2.4 of the MCA model), instead for *bla*_NDM_ the situation is the opposite (5.7 in the MCA model compared to the 4.6 of the ACA model).

#### 2.2 Supplementary Tables

**Supplementary Tables S1**. Bacterial isolates used in this study

| **Sample number** | **Species**  **(MALDI-TOF)** | **Sample source** | **Collection date** | **Carbapene-mase gene carried** | **AMCA classification** | **Conc. (cp/µl)** |
| --- | --- | --- | --- | --- | --- | --- |
| CPO001 | *Acinetobacter baumannii* | Bronchoalveolar lavage | 26/02/2015 | oxa23 | neg | 0.00E+00 |
| CPO002 | *Klebsiella pneumoniae* | Urine | 15/04/2013 | neg | neg | 0.00E+00 |
| CPO003 | *Klebsiella pneumoniae* | Rectal swab | 29/05/2018 | neg | neg | 0.00E+00 |
| CPO004 | *Enterobacter cloacae* | Right leg tissue | 13/07/2018 | neg | neg | 0.00E+00 |
| CPO005 | *Escherichia coli* | Sputum | 10/10/2015 | neg | neg | 0.00E+00 |
| CPO006 | *Escherichia coli* | Urine | 04/01/2016 | neg | neg | 0.00E+00 |
| CPO007 | *Klebsiella pneumoniae* | Urine | 29/02/2016 | neg | neg | 0.00E+00 |
| CPO008 | *Citrobacter freundii* | Urine | 09/03/2016 | neg | neg | 0.00E+00 |
| CPO009 | *Klebsiella pneumoniae* | Urine | 10/07/2016 | neg | neg | 0.00E+00 |
| CPO010 | *Enterobacter cloacae* | Bronchoalveolar lavage | 15/08/2016 | neg | neg | 0.00E+00 |
| CPO011 | *Klebsiella pneumoniae* | Perineum | 05/10/2016 | neg | neg | 0.00E+00 |
| CPO012 | *Klebsiella pneumoniae* | Right leg tissue | 23/10/2016 | neg | neg | 0.00E+00 |
| CPO013 | *Klebsiella pneumoniae* | Rectal swab | 26/12/2016 | neg | neg | 0.00E+00 |
| CPO014 | *Escherichia coli* | Urine | 18/06/2017 | neg | neg | 0.00E+00 |
| CPO015 | *Klebsiella pneumoniae* | Urine | 18/06/2017 | neg | neg | 0.00E+00 |
| CPO016 | *Klebsiella pneumoniae* | Urine | 05/08/2017 | neg | neg | 0.00E+00 |
| CPO017 | *Enterobacter cloacae* | Sputum | 18/08/2017 | neg | neg | 0.00E+00 |
| CPO018 | *Escherichia coli* | Urine | 18/08/2017 | neg | neg | 0.00E+00 |
| CPO019 | *Klebsiella pneumoniae* | Urine | 27/10/2017 | neg | neg | 0.00E+00 |
| CPO020 | *Klebsiella pneumoniae* | Urine | 05/01/2018 | neg | neg | 0.00E+00 |
| CPO021 | *Enterobacter cloacae* | Wound swab | 14/11/2017 | neg | neg | 0.00E+00 |
| CPO022 | *Escherichia coli* | Rectal swab | 22/01/2018 | neg | neg | 0.00E+00 |
| CPO023 | *Escherichia coli* | Urine | 22/01/2018 | neg | neg | 0.00E+00 |
| CPO024 | *Klebsiella pneumoniae* | Urine | 26/01/2018 | neg | neg | 0.00E+00 |
| CPO025 | *Enterobacter spp* | Rectal swab | 16/05/2019 | neg | neg | 0.00E+00 |
| CPO026 | *Escherichia coli* | Rectal swab | 16/05/2019 | neg | neg | 0.00E+00 |
| CPO027 | *Enterobacter cloacae* | Rectal swab | 12/05/2018 | neg | neg | 0.00E+00 |
| CPO028 | *Escherichia coli* | Rectal swab | 22/04/2018 | neg | neg | 0.00E+00 |
| CPO029 | *Klebsiella pneumoniae* | Rectal swab | 07/01/2019 | neg | neg | 0.00E+00 |
| CPO030 | *Pseudomonas aeruginosa* | Rectal swab | 12/01/2019 | neg | neg | 0.00E+00 |
| CPO031 | *Escherichia coli* | Rectal swab | 11/02/2019 | neg | neg | 0.00E+00 |
| CPO032 | *Acinetobacter baumannii* | Rectal swab | 13/03/2019 | neg | neg | 0.00E+00 |
| CPO033 | *Pseudomonas aeruginosa* | Throat swab | 31/03/2015 | imp | imp | 2.42E+03 |
| CPO034 | *Escherichia coli* | Rectal swab | 12/05/2018 | imp | imp | 1.64E+04 |
| CPO035 | *Klebsiella pneumoniae* | Rectal swab | 18/05/2018 | imp | imp | 2.06E+03 |
| CPO036 | *Enterobacter cloacae* | Rectal swab | 23/06/2018 | imp | imp | 9.88E+01 |
| CPO037 | *Klebsiella pneumoniae* | Rectal swab | 03/07/2018 | imp | imp | 8.00E+02 |
| CPO038 | *Escherichia coli* | Rectal swab | 13/01/2019 | imp | imp | 3.24E+04 |
| CPO039 | *Klebsiella pneumoniae* | Rectal swab | 14/01/2019 | imp | imp | 3.28E+04 |
| CPO040 | *Escherichia coli* | Rectal swab | 30/01/2019 | imp | imp | 1.36E+04 |
| CPO041 | *Enterobacter cloacae* | Rectal swab | 27/07/2019 | imp | imp | 2.11E+02 |
| CPO042 | *Klebsiella pneumoniae* | Rectal swab | 24/07/2019 | imp | imp | 9.11E+03 |
| CPO043 | *Klebsiella pneumoniae* | Rectal swab | 29/07/2019 | imp | imp | 3.69E+03 |
| CPO044 | *Enterobacter cloacae* | Rectal swab | 26/08/2019 | imp | imp | 6.58E+03 |
| CPO045 | *Enterobacter cloacae* | Rectal swab | 18/08/2019 | imp | imp | 1.67E+04 |
| CPO046 | *Klebsiella pneumoniae* | Rectal swab | 06/05/2019 | imp | imp | 2.59E+04 |
| CPO047 | *Enterobacter cloacae* | Rectal swab | 09/05/2019 | imp | imp | 1.46E+02 |
| CPO048 | *Enterobacter cloacae* | Rectal swab | 11/05/2019 | imp | imp | 1.75E+04 |
| CPO049 | *Enterobacter spp* | Rectal swab | 13/06/2019 | imp | imp | 3.27E+04 |
| CPO050 | *Escherichia coli* | Rectal swab | 20/06/2019 | imp | imp | 9.15E+03 |
| CPO051 | *Enterobacter cloacae* | Rectal swab | 09/09/2017 | imp | imp | 2.32E+04 |
| CPO052 | *Klebsiella pneumoniae* | Rectal swab | 03/10/2017 | imp | imp | 4.27E+01 |
| CPO053 | *Enterobacter cloacae* | Rectal swab | 05/10/2017 | imp | imp | 2.18E+04 |
| CPO054 | *Enterobacter spp* | Rectal swab | 11/10/2017 | imp | imp | 1.60E+04 |
| CPO055 | *Citrobacter freundii* | Rectal swab | 28/10/2017 | imp | imp | 5.55E+02 |
| CPO056 | *Klebsiella pneumoniae* | Rectal swab | 22/01/2018 | imp | imp | 1.20E+03 |
| CPO057 | *Klebsiella pneumoniae* | Rectal swab | 28/01/2018 | imp | imp | 9.89E+02 |
| CPO058 | *Enterobacter cloacae* | Urine | 06/02/2018 | imp | imp | 7.53E+01 |
| CPO059 | *Enterobacter spp* | Rectal swab | 21/02/2018 | imp | imp | 7.92E+02 |
| CPO060 | *Klebsiella pneumoniae* | Rectal swab | 10/09/2018 | imp | imp | 1.89E+02 |
| CPO061 | *Enterobacter cloacae* | Rectal swab | 07/09/2018 | imp | imp | 1.83E+02 |
| CPO062 | *Escherichia hermannii* | Rectal swab | 20/11/2018 | imp | imp | 1.47E+02 |
| CPO063 | *Escherichia coli* | Rectal swab | 21/02/2018 | imp | imp | 1.05E+03 |
| CPO064 | *Enterobacter cloacae* | Rectal swab | 28/01/2018 | imp | imp | 1.20E+02 |
| CPO065 | *Klebsiella pneumoniae* | Rectal swab | 02/04/2018 | imp | imp | 3.48E+04 |
| CPO066 | *Klebsiella pneumoniae* | Rectal swab | 22/04/2018 | imp | imp | 3.45E+04 |
| CPO067 | *Enterobacter cloacae* | Rectal swab | 18/08/2018 | imp | imp | 1.05E+02 |
| CPO068 | *Klebsiella pneumoniae* | Rectal swab | 07/01/2019 | imp | imp | 1.72E+02 |
| CPO069 | *Pseudomonas aeruginosa* | Rectal swab | 12/01/2019 | imp | imp | 4.95E+03 |
| CPO070 | *Enterobacter cloacae* | Rectal swab | 11/02/2019 | imp | imp | 2.41E+02 |
| CPO071 | *Klebsiella pneumoniae* | Rectal swab | 13/03/2019 | imp | imp | 4.20E+02 |
| CPO072 | *Escherichia hermannii* | Rectal swab | 27/05/2019 | imp | imp | 3.01E+02 |
| CPO073 | *Enterobacter spp* | Rectal swab | 08/05/2019 | imp | imp | 1.32E+02 |
| CPO074 | *Klebsiella pneumoniae* | Swab | 11/05/2019 | imp | imp | 3.45E+01 |
| CPO075 | *Enterobacter cloacae* | Rectal swab | 16/06/2016 | imp | imp | 5.43E+01 |
| CPO076 | *Enterobacter cloacae* | Rectal swab | 16/05/2019 | imp | imp | 2.41E+04 |
| CPO077 | *Enterobacter cloacae* | Rectal swab | 12/05/2018 | imp | imp | 1.51E+04 |
| CPO078 | *Klebsiella pneumoniae* | Wound swab | 08/10/2012 | kpc | kpc | 5.83E+03 |
| CPO079 | *Citrobacter spp* | Rectal Swab | 01/10/2017 | kpc | kpc | 9.73E+03 |
| CPO080 | *Klebsiella pneumoniae* | Urine | 22/03/2014 | kpc | kpc | 1.22E+04 |
| CPO081 | *Klebsiella pneumoniae* | Rectal Swab | 10/09/2017 | kpc | kpc | 3.48E+03 |
| CPO082 | *Citrobacter spp* | Rectal Swab | 15/10/2017 | kpc | kpc | 3.45E+03 |
| CPO083 | *Klebsiella pneumoniae* | Urine | 08/04/2015 | kpc | kpc | 7.61E+03 |
| CPO084 | *Serratia marcescens* | Rectal Swab | 10/10/2017 | kpc | kpc | 8.66E+03 |
| CPO085 | *Klebsiella pneumoniae* | Rectal Swab | 11/09/2017 | kpc | kpc | 3.98E+03 |
| CPO086 | *Klebsiella pneumoniae* | Rectal Swab | 11/09/2017 | kpc | kpc | 1.82E+04 |
| CPO087 | *Escherichia coli* | Rectal Swab | 01/08/2016 | ndm | ndm | 4.23E+03 |
| CPO088 | *Klebsiella pneumoniae* | Urine | 25/12/2015 | ndm | ndm | 5.39E+03 |
| CPO089 | *Klebsiella pneumoniae* | Rectal Swab | 18/12/2015 | ndm | ndm | 6.47E+03 |
| CPO090 | *Escherichia coli* | Bone (Tibia) | 14/01/2015 | ndm | ndm | 3.69E+03 |
| CPO091 | *Klebsiella pneumoniae* | Throat swab | 02/08/2016 | ndm | ndm | 3.66E+03 |
| CPO092 | *Klebsiella pneumoniae* | Rectal Swab | 18/04/2015 | ndm | ndm | 1.19E+04 |
| CPO093 | *Klebsiella pneumoniae* | Urine | 23/04/2015 | ndm | ndm | 4.26E+03 |
| CPO094 | *Klebsiella pneumoniae* | Rectal Swab | 25/04/2015 | ndm | ndm | 2.22E+03 |
| CPO095 | *Proteus mirabilis* | Urine | 07/02/2014 | ndm | ndm | 1.40E+03 |
| CPO096 | *Klebsiella pneumoniae* | Rectal Swab | 04/12/2016 | ndm | ndm | 2.63E+03 |
| CPO097 | *Klebsiella pneumoniae* | Mouth Swab | 29/01/2015 | ndm | ndm | 2.48E+03 |
| CPO098 | *Klebsiella pneumoniae* | Rectal Swab | 20/04/2015 | ndm | ndm | 2.39E+03 |
| CPO099 | *Klebsiella pneumoniae* | Perinrum swab | 10/05/2015 | ndm | ndm | 1.00E+04 |
| CPO100 | *Escherichia coli* | Vaginal swab | 05/03/2015 | ndm | ndm | 1.40E+04 |
| CPO101 | *Escherichia coli* | Perinrum swab | 09/04/2015 | ndm | ndm | 9.82E+03 |
| CPO102 | *Klebsiella pneumoniae* | Perinrum swab | 09/12/2014 | ndm | ndm | 1.56E+04 |
| CPO103 | *Klebsiella pneumoniae* | Rectal Swab | 20/04/2015 | ndm | ndm | 1.66E+04 |
| CPO104 | *Klebsiella pneumoniae* | Rectal Swab | 15/04/2015 | ndm | ndm | 1.06E+04 |
| CPO105 | *Escherichia coli* | Faeces | 23/11/2015 | ndm | ndm | 9.28E+03 |
| CPO106 | *Escherichia coli* | Rectal Swab | 26/12/2015 | ndm | ndm | 7.51E+03 |
| CPO107 | *Klebsiella pneumoniae* | Perinrum swab | 07/05/2015 | ndm | ndm | 6.29E+03 |
| CPO108 | *Klebsiella pneumoniae* | Rectal Swab | 20/04/2015 | ndm | ndm | 1.85E+04 |
| CPO109 | *Klebsiella pneumoniae* | Sputum | 23/03/2018 | ndm | ndm | 9.74E+03 |
| CPO110 | *Klebsiella pneumoniae* | Rectal Swab | 29/04/2015 | ndm | ndm | 1.27E+04 |
| CPO111 | *Klebsiella pneumoniae* | Perinrum swab | 07/05/2015 | ndm | ndm | 1.05E+04 |
| CPO112 | *Klebsiella pneumoniae* | Catheter Urine | 02/07/2014 | ndm | ndm | 1.21E+04 |
| CPO113 | *Klebsiella pneumoniae* | Perinrum swab | 09/12/2014 | ndm | ndm | 1.50E+04 |
| CPO114 | *Klebsiella pneumoniae* | Urine | 08/03/2015 | ndm | ndm | 8.01E+03 |
| CPO115 | *Klebsiella pneumoniae* | Urine | 10/04/2015 | ndm | ndm | 2.06E+04 |
| CPO116 | *Klebsiella pneumoniae* | Perinrum swab | 19/04/2015 | ndm | ndm | 1.46E+04 |
| CPO117 | *Klebsiella pneumoniae* | Rectal Swab | 18/04/2015 | ndm | ndm | 2.83E+04 |
| CPO118 | *Klebsiella pneumoniae* | Rectal Swab | 17/05/2015 | ndm | ndm | 9.15E+03 |
| CPO119 | *Klebsiella pneumoniae* | Urine | 23/04/2015 | ndm | ndm | 2.22E+04 |
| CPO120 | *Klebsiella pneumoniae* | Rectal Swab | 25/04/2015 | ndm | ndm | 1.41E+04 |
| CPO121 | *Klebsiella pneumoniae* | Perineum swab | 10/05/2015 | ndm | ndm | 8.60E+03 |
| CPO122 | *Klebsiella pneumoniae* | Rectal Swab | 03/10/2015 | ndm | ndm | 2.08E+04 |
| CPO123 | *Klebsiella pneumoniae* | Rectal Swab | 16/08/2015 | ndm | ndm | 1.96E+04 |
| CPO124 | *Klebsiella pneumoniae* | Wound swab | 19/04/2015 | ndm | ndm | 6.44E+03 |
| CPO125 | *Klebsiella pneumoniae* | Rectal Swab | 16/09/2015 | ndm | ndm | 1.68E+04 |
| CPO126 | *Klebsiella pneumoniae* | Rectal Swab | 07/06/2015 | ndm | ndm | 1.64E+04 |
| CPO127 | *Klebsiella pneumoniae* | Rectal Swab | 07/07/2015 | ndm | ndm | 2.29E+04 |
| CPO128 | *Klebsiella pneumoniae* | Rectal Swab | 03/06/2015 | ndm | ndm | 6.54E+03 |
| CPO129 | *Klebsiella pneumoniae* | Rectal Swab | 20/05/2015 | ndm | ndm | 1.18E+04 |
| CPO130 | *Klebsiella pneumoniae* | Rectal Swab | 10/08/2015 | ndm | ndm | 7.64E+03 |
| CPO131 | *Klebsiella pneumoniae* | Perineum swab | 12/07/2015 | ndm | ndm | 6.86E+03 |
| CPO132 | *Klebsiella pneumoniae* | Rectal Swab | 21/07/2015 | ndm | ndm | 2.40E+04 |
| CPO133 | *Escherichia coli* | Rectal swab | 29/06/2018 | ndm | ndm | 1.02E+04 |
| CPO134 | *Klebsiella pneumoniae* | Blood culture | 23/09/2016 | ndm | ndm | 3.14E+04 |
| CPO135 | *Enterobacter cloacae* | Rectal swab | 24/06/2018 | ndm | ndm & kpc | 5.75E+04 |
| CPO136 | *Escherichia coli* | Urine | 12/03/2019 | ndm | ndm | 1.83E+04 |
| CPO137 | *Escherichia coli* | Rectal swab | 28/01/2019 | ndm | ndm | 1.29E+04 |
| CPO138 | *Klebsiella pneumoniae* | Rectal swab | 23/02/2019 | ndm | ndm | 2.41E+04 |
| CPO139 | *Klebsiella pneumoniae* | Rectal swab | 25/03/2019 | ndm | ndm | 5.85E+04 |
| CPO140 | *Enterobacter cloacae* | Rectal swab | 03/04/2019 | ndm | ndm | 1.24E+04 |
| CPO141 | *Enterobacter cloacae* | Rectal swab | 21/01/2019 | ndm | ndm | 8.38E+03 |
| CPO142 | *Citrobacter freundii* | Abdomen | 08/04/2019 | ndm | ndm | 1.34E+04 |
| CPO143 | *Klebsiella pneumoniae* | Urine | 23/04/2015 | ndm | ndm | 1.11E+04 |
| CPO144 | *Klebsiella pneumoniae* | Right leg tissue | 29/07/2015 | ndm | ndm | 7.75E+03 |
| CPO145 | *Klebsiella pneumoniae* | Abdomen | 22/09/2016 | ndm | ndm | 1.55E+04 |
| CPO146 | *Escherichia coli* | Urine | 04/05/2017 | ndm | ndm | 1.73E+04 |
| CPO147 | *Escherichia coli* | Catheter urine | 06/06/2019 | ndm | ndm | 6.45E+03 |
| CPO148 | *Escherichia coli* | Rectal swab | 12/01/2019 | ndm | ndm | 1.13E+05 |
| CPO149 | *Escherichia coli* | Rectal swab | 27/05/2019 | ndm | ndm | 2.44E+05 |
| CPO150 | *Enterobacter cloacae* | Rectal swab | 16/06/2016 | ndm | ndm | 2.72E+06 |
| CPO151 | *Klebsiella pneumoniae* | Rectal swab | 16/05/2019 | ndm | ndm | 7.59E+04 |
| CPO152 | *Enterobacter cloacae* | Rectal swab | 12/05/2018 | ndm | ndm | 2.48E+06 |
| CPO153 | *Enterobacter cloacae* | Rectal swab | 22/04/2018 | ndm | ndm | 1.80E+06 |
| CPO154 | *Escherichia coli* | Rectal swab | 18/08/2018 | ndm | ndm | 3.59E+05 |
| CPO155 | *Klebsiella pneumoniae* | Rectal swab | 08/05/2019 | ndm | ndm | 1.87E+06 |
| CPO156 | *Enterobacter cloacae* | Rectal swab | 12/05/2018 | ndm | ndm | 1.11E+05 |
| CPO157 | *Klebsiella pneumoniae* | Rectal swab | 07/01/2019 | ndm | ndm | 1.54E+06 |
| CPO158 | *Klebsiella pneumoniae* | Rectal swab | 12/01/2019 | ndm | ndm | 2.27E+06 |
| CPO159 | *Klebsiella pneumoniae* | Rectal swab | 13/03/2019 | ndm | ndm | 3.15E+04 |
| CPO160 | *Klebsiella pneumoniae* | Rectal swab | 27/05/2019 | ndm | ndm | 9.98E+04 |
| CPO161 | *Escherichia coli* | Rectal Swab | 01/12/2014 | ndm & oxa48 | ndm & oxa48 | 1.70E+04 |
| CPO162 | *Escherichia coli* | Wound swab | 14/03/2014 | oxa48 | oxa48 | 2.45E+04 |
| CPO163 | *Escherichia coli* | Rectal Swab | 20/10/2017 | oxa48 | oxa48 | 3.76E+03 |
| CPO164 | *Citrobacter freundii* | Rectal Swab | 06/01/2016 | oxa48 | oxa48 | 5.56E+03 |
| CPO165 | *Escherichia coli* | Urine | 04/04/2015 | oxa48 | oxa48 | 2.48E+03 |
| CPO166 | *Klebsiella pneumoniae* | Rectal Swab | 07/01/2016 | oxa48 | oxa48 | 1.50E+04 |
| CPO167 | *Escherichia coli* | Wound swab | 25/11/2012 | oxa48 | oxa48 | 2.22E+03 |
| CPO168 | *Escherichia coli* | Blood culture | 21/07/2013 | oxa48 | oxa48 | 1.17E+04 |
| CPO169 | *Serratia marcescens* | Bone (Tibia) | 28/01/2015 | oxa48 | oxa48 | 6.25E+03 |
| CPO170 | *Citrobacter freundii* | Rectal Swab | 13/12/2015 | oxa48 | oxa48 | 3.23E+03 |
| CPO171 | *Klebsiella pneumoniae* | Urine | 02/07/2014 | oxa48 | oxa48 | 1.46E+03 |
| CPO172 | *Klebsiella pneumoniae* | Drain Fluid Abdomen | 22/01/2015 | oxa48 | oxa48 | 4.55E+03 |
| CPO173 | *Escherichia coli* | Urine | 04/04/2015 | oxa48 | oxa48 | 2.25E+04 |
| CPO174 | *Escherichia coli* | Rectal swab | 28/06/2018 | oxa48 | oxa48 | 1.16E+04 |
| CPO175 | *Klebsiella pneumoniae* | Urine | 08/05/2018 | oxa48 | oxa48 | 3.36E+04 |
| CPO176 | *Klebsiella pneumoniae* | Blood culture | 13/06/2018 | oxa48 | oxa48 | 2.77E+04 |
| CPO177 | *Klebsiella pneumoniae* | Blood culture | 15/10/2018 | oxa48 | oxa48 | 3.40E+04 |
| CPO178 | *Klebsiella pneumoniae* | Blood culture | 21/02/2015 | oxa48 | oxa48 | 8.70E+03 |
| CPO179 | *Citrobacter freundii* | Rectal swab | 14/01/2018 | oxa48 | oxa48 | 4.21E+04 |
| CPO180 | *Citrobacter freundii* | Urine | 16/01/2018 | oxa48 | oxa48 | 3.12E+04 |
| CPO181 | *Klebsiella pneumoniae* | Rectal swab | 04/03/2019 | oxa48 | oxa48 | 6.24E+04 |
| CPO182 | *Klebsiella pneumoniae* | Urine | 30/08/2018 | oxa48 | oxa48 | 2.14E+04 |
| CPO183 | *Klebsiella pneumoniae* | Urine | 23/04/2019 | oxa48 | oxa48 | 8.64E+04 |
| CPO184 | *Klebsiella pneumoniae* | Urine | 01/06/2015 | oxa48 | oxa48 | 2.82E+05 |
| CPO185 | *Escherichia coli* | Wound swab | 08/07/2015 | oxa48 | oxa48 | 1.58E+04 |
| CPO186 | *Klebsiella pneumoniae* | Urine | 04/01/2016 | oxa48 | oxa48 | 9.71E+04 |
| CPO187 | *Citrobacter amalonaticus* | Rectal swab | 18/01/2016 | oxa48 | oxa48 | 3.38E+04 |
| CPO188 | *Klebsiella pneumoniae* | Urine | 08/02/2016 | oxa48 | oxa48 | 8.94E+05 |
| CPO189 | *Klebsiella pneumoniae* | Urine | 08/02/2016 | oxa48 | oxa48 | 4.47E+05 |
| CPO190 | *Klebsiella pneumoniae* | Wound swab | 01/07/2016 | oxa48 | oxa48 | 5.80E+05 |
| CPO191 | *Klebsiella pneumoniae* | Pleural fluid | 10/07/2016 | oxa48 | oxa48 | 7.23E+05 |
| CPO192 | *Escherichia coli* | Rectal swab | 15/08/2016 | oxa48 | oxa48 | 4.37E+04 |
| CPO193 | *Escherichia coli* | Urine | 26/08/2016 | oxa48 | oxa48 | 2.14E+04 |
| CPO194 | *Klebsiella pneumoniae* | Urine | 22/10/2016 | oxa48 | oxa48 | 1.35E+05 |
| CPO195 | *Escherichia coli* | Urine | 27/11/2016 | oxa48 | oxa48 | 1.11E+04 |
| CPO196 | *Klebsiella pneumoniae* | Rectal swab | 18/03/2017 | oxa48 | oxa48 | 5.28E+05 |
| CPO197 | *Escherichia coli* | Wound swab | 18/04/2017 | oxa48 | oxa48 | 9.71E+03 |
| CPO198 | *Citrobacter freundii* | Urine | 05/05/2017 | oxa48 | oxa48 | 5.71E+03 |
| CPO199 | *Klebsiella pneumoniae* | Wound swab | 06/08/2017 | oxa48 | oxa48 | 1.29E+04 |
| CPO200 | *Klebsiella pneumoniae* | Abdomen | 17/01/2018 | oxa48 | oxa48 | 1.73E+03 |
| CPO201 | *Klebsiella pneumoniae* | Rectal swab | 22/01/2018 | oxa48 | oxa48 | 1.78E+03 |
| CPO202 | *Klebsiella pneumoniae* | Urine | 27/01/2018 | oxa48 | oxa48 | 1.26E+05 |
| CPO203 | *Escherichia coli* | Rectal swab | 22/04/2018 | oxa48 | oxa48 | 1.02E+06 |
| CPO204 | *Citrobacter freundii* | Rectal swab | 18/08/2018 | oxa48 | oxa48 | 5.77E+05 |
| CPO205 | *Escherichia coli* | Rectal swab | 07/01/2019 | oxa48 | oxa48 | 2.69E+05 |
| CPO206 | *Escherichia coli* | Rectal swab | 12/01/2019 | oxa48 | oxa48 | 1.78E+04 |
| CPO207 | *Citrobacter freundii* | Rectal swab | 11/02/2019 | oxa48 | oxa48 | 2.92E+06 |
| CPO208 | *Escherichia coli* | Rectal swab | 13/03/2019 | oxa48 | oxa48 | 1.57E+04 |
| CPO209 | *Klebsiella pneumoniae* | Rectal swab | 27/05/2019 | oxa48 | oxa48 | 2.45E+06 |
| CPO210 | *Klebsiella pneumoniae* | Rectal swab | 08/05/2019 | oxa48 | oxa48 | 2.03E+06 |
| CPO211 | *Klebsiella pneumoniae* | Rectal swab | 11/05/2019 | oxa48 | oxa48 | 2.41E+05 |
| CPO212 | *Klebsiella pneumoniae* | Rectal swab | 16/06/2016 | oxa48 | oxa48 | 3.61E+05 |
| CPO213 | *Escherichia coli* | Rectal swab | 16/05/2019 | oxa48 | oxa48 | 3.29E+04 |
| CPO214 | *Klebsiella pneumoniae* | Rectal swab | 12/05/2018 | oxa48 | oxa48 | 1.21E+06 |
| CPO215 | *Klebsiella pneumoniae* | Rectal swab | 22/04/2018 | oxa48 | oxa48 | 1.57E+06 |
| CPO216 | *Klebsiella pneumoniae* | Rectal swab | 18/08/2018 | oxa48 | oxa48 | 4.18E+05 |
| CPO217 | *Klebsiella pneumoniae* | Rectal swab | 07/01/2019 | oxa48 | oxa48 | 2.43E+06 |
| CPO218 | *Escherichia coli* | Rectal swab | 12/01/2019 | oxa48 | oxa48 | 1.80E+05 |
| CPO219 | *Escherichia coli* | Rectal swab | 11/02/2019 | oxa48 | oxa48 | 2.78E+03 |
| CPO220 | *Klebsiella pneumoniae* | Rectal swab | 13/03/2019 | oxa48 | oxa48 | 3.75E+05 |
| CPO221 | *Escherichia coli* | Rectal swab | 27/05/2019 | oxa48 | oxa48 | 5.40E+06 |
| CPO222 | *Klebsiella pneumoniae* | Rectal swab | 08/05/2019 | oxa48 | oxa48 | 9.42E+03 |
| CPO223 | *Klebsiella pneumoniae* | Rectal swab | 11/05/2019 | oxa48 | oxa48 | 3.71E+05 |
| CPO224 | *Citrobacter freundii* | Rectal swab | 16/06/2016 | oxa48 | oxa48 | 7.02E+05 |
| CPO225 | *Escherichia coli* | Rectal swab | 12/05/2018 | oxa48 | oxa48 | 8.26E+05 |
| CPO226 | *Klebsiella pneumoniae* | Rectal swab | 22/04/2018 | oxa48 | oxa48 | 7.85E+05 |
| CPO227 | *Escherichia coli* | Rectal swab | 18/08/2018 | oxa48 | oxa48 | 4.46E+05 |
| CPO228 | *Citrobacter freundii* | Rectal swab | 07/01/2019 | oxa48 | oxa48 | 5.04E+05 |
| CPO229 | *Klebsiella pneumoniae* | Rectal swab | 11/02/2019 | oxa48 | oxa48 | 2.57E+05 |
| CPO230 | *Klebsiella pneumoniae* | Rectal swab | 13/03/2019 | oxa48 | oxa48 | 4.65E+05 |
| CPO231 | *Klebsiella pneumoniae* | Rectal swab | 08/05/2019 | oxa48 | oxa48 | 4.16E+05 |
| CPO232 | *Klebsiella pneumoniae* | Rectal swab | 11/05/2019 | oxa48 | oxa48 | 1.03E+05 |
| CPO233 | *Klebsiella pneumoniae* | Rectal swab | 07/01/2019 | oxa48 | oxa48 | 2.01E+06 |
| CPO234 | *Klebsiella pneumoniae* | Rectal swab | 12/01/2019 | oxa48 | oxa48 | 6.13E+04 |
| CPO235 | *Escherichia coli* | Rectal swab | 11/02/2019 | oxa48 | oxa48 | 2.54E+05 |
| CPO236 | *Klebsiella pneumoniae* | Rectal swab | 13/03/2019 | oxa48 | oxa48 | 2.26E+05 |
| CPO237 | *Escherichia coli* | Rectal swab | 27/05/2019 | oxa48 | oxa48 | 2.22E+05 |
| CPO238 | *Escherichia coli* | Rectal swab | 11/05/2019 | oxa48 | oxa48 | 8.02E+05 |
| CPO239 | *Klebsiella pneumoniae* | Rectal swab | 16/06/2016 | oxa48 | oxa48 | 2.90E+05 |
| CPO240 | *Klebsiella pneumoniae* | Rectal swab | 16/05/2019 | oxa48 | oxa48 | 2.93E+05 |
| CPO241 | *Escherichia coli* | Rectal swab | 22/04/2018 | oxa48 | oxa48 | 5.33E+05 |
| CPO242 | *Enterobacter cloacae* | Rectal swab | 18/08/2018 | oxa48 | oxa48 | 8.86E+04 |
| CPO243 | *Klebsiella pneumoniae* | Rectal swab | 16/06/2016 | oxa48 | oxa48 | 4.46E+06 |
| CPO244 | *Klebsiella pneumoniae* | Rectal swab | 18/08/2018 | oxa48 | oxa48 | 2.11E+06 |
| CPO245 | *Enterobacter spp* | Rectal swab | 27/05/2019 | oxa48 | oxa48 | 4.32E+04 |
| CPO246 | *Pseudomonas aeruginosa* | Wound swab | 25/03/2015 | vim | vim | 2.28E+03 |
| CPO247 | *Citrobacter freundii* | Rectal Swab | 02/04/2016 | vim | vim | 1.33E+04 |
| CPO248 | *Enterobacter cloacae* | Bone (Tibia) | 14/01/2015 | vim | vim | 5.66E+03 |
| CPO249 | *Pseudomonas aeruginosa* | Sputum | 01/11/2013 | vim | vim | 5.19E+03 |
| CPO250 | *Enterobacter cloacae* | Bone (Tibia) | 14/01/2015 | vim | vim | 2.95E+04 |
| CPO251 | *Klebsiella pneumoniae* | Rectal swab | 11/02/2019 | vim | vim | 1.49E+05 |
| CPO252 | *Klebsiella pneumoniae* | Rectal swab | 08/05/2019 | vim | vim | 1.90E+05 |
| CPO253 | *Klebsiella pneumoniae* | Rectal swab | 11/05/2019 | vim | vim | 1.39E+05 |
